# Germline polygenic score for prostate cancer aggressiveness

**DOI:** 10.64898/2026.05.07.26352488

**Authors:** George Xu, Roshan Karunamuni, Anna M Dornisch, Charles A Brunette, Morgan E Danowski, Heena Desai, Daniel Dochtermann, Isla P Garraway, Richard L Hauger, Adam S Kibel, Julie A Lynch, Saiju Pyarajan, Brent S Rose, Craig C Teerlink, Ole A Andreassen, Anders M Dale, Jenny L Donovan, Freddie Hamdy, Linda Kachuri, Athene Lane, Richard M Martin, Ian G Mills, David E Neal, Emma L Turner, John S Witte, Johanna Schleutker, Nora Pashayan, Jyotsna Batra, Australian Prostate Cancer BioResource (APCB), Børge G Nordestgaard, Robert J Hamilton, Alicja Wolk, Demetrius Albanes, Joshua Atkins, William J Blot, Lorelei A Mucci, Sune F Nielsen, Olivier Cussenot, Sonja I Berndt, Stella Koutros, Karina Dalsgaard Sørensen, Cezary Cybulski, Florence Menegaux, Jong Y Park, Robert J MacInnis, Barry S Rosenstein, Yong-Jie Lu, Stephen Watya, Ana Vega, NC-LA PCaP Investigators, The IMPACT Study Steering Committee and Collaborators, Manolis Kogevinas, Fredrik Wiklund, Anna Plym, Manuel R Teixeira, Luc Multigner, Robin J Leach, Hermann Brenner, Esther M John, Radka Kaneva, Christopher J Logothetis, Susan L Neuhausen, Piet Ost, Azad Razack, Jay H Fowke, Marija Gamulin, Nawaid Usmani, Frank Claessens, Jose Esteban Castelao, Gyorgy Petrovics, Marie-Élise Parent, Jennifer J Hu, Wei Zheng, The Profile Study Steering Committee, UKGPCS collaborators, Zsofia Kote-Jarai, Rosalind A Eeles, The PRACTICAL Consortium, VA Million Veteran Program, Kara N Maxwell, Jason L Vassy, Tyler M Seibert

**Affiliations:** Research Service, VA San Diego Healthcare System, San Diego, CA, USA; Department of Radiation Medicine and Applied Sciences, University of California San Diego, La Jolla, CA, USA; VA Boston Healthcare System, Boston, MA, USA; Department of Medicine, Harvard Medical School, Boston, MA, USA; Department of Medicine, Hematology-Oncology, Perelman School of Medicine, University of Pennsylvania, Philadelphia, PA; Center for Data and Computational Sciences, VA Boston Healthcare System, Boston, MA, USA; Division of Urology, VA Greater Los Angeles Healthcare System, Los Angeles, CA, USA; Department of Urology, David Geffen School of Medicine at UCLA, Los Angeles, CA, USA; Center for Behavior Genetics of Aging, School of Medicine, UC San Diego, La Jolla, CA, USA; Harvard Medical School, Boston, MA, USA; Department of Urology, Mass General Brigham, Harvard Medical School, Boston, MA, USA; VA Informatics and Computing Infrastructure, VA Salt Lake City Healthcare System, Salt Lake City, UT, USA; Department of Internal Medicine, University of Utah School of Medicine, Salt Lake City, CA, USA; Center for Precision Psychiatry, Oslo University Hospital and University of Oslo, Oslo, Norway; Center for Multimodal Imaging and Genetics, University of California, San Diego, La Jolla, CA, USA; Department of Radiology, University of California, San Diego, La Jolla, CA, USA; Department of Neurosciences, University of California, San Diego, La Jolla, CA, USA; Halıcıoğlu Data Science Institute, University of California San Diego, La Jolla, California, USA; NORMENT, KG Jebsen Centre, Oslo University Hospital and University of Oslo, Oslo, Norway; Population Health Sciences, Bristol Medical School, University of Bristol, Bristol, UK; Nuffield Department of Surgical Sciences, University of Oxford, Oxford, UK; Cancer Research UK Oxford Centre, Old Road Campus Research Building, Oxford OX2 7DQ, UK; Department of Epidemiology and Population Health, Stanford University, Stanford, CA, USA; National Institute for Health Research (NIHR) Bristol Biomedical Research Centre, University Hospitals Bristol and Weston NHS Foundation Trust and University of Bristol, Bristol, UK; University of Cambridge, Department of Oncology, Box 279, Addenbrooke’s Hospital, Hills Road, Cambridge CB2 0QQ, UK; Cancer Research UK, Cambridge Research Institute, Li Ka Shing Centre, Cambridge, CB2 0RE, UK; Institute of Biomedicine, University of Turku, Finland; Department of Genomics, Laboratory Division, Turku University Hospital, PO Box 52, 20521 Turku, Finland; Department of Public Health & Primary Care, Strangeways Research Laboratory, Worts Causeway, Cambridge, CB1 8RN; School of Biomedical Science, Faculty of Health Sciences and Medicine, Bond University, Gold Coast, Queensland, Australia; Centre for Genomics and Personalised Health, School of Biomedical Sciences, Faculty of Health, Queensland University of Technology, Brisbane, Queensland, Australia; Translational Research Institute, Brisbane, Queensland 4102, Australia; Centre for Genomics and Personalised Health, Queensland University of Technology, Brisbane; Prostate Cancer Research Program, Monash University, Melbourne; Dame Roma Mitchell Cancer Centre, University of Adelaide, Adelaide; Chris O’Brien Lifehouse and The Kinghorn Cancer Centre, Sydney, Australia; Faculty of Health and Medical Sciences, University of Copenhagen, 2200 Copenhagen, Denmark; Department of Clinical Biochemistry, Herlev and Gentofte Hospital, Copenhagen University Hospital, Herlev, 2200 Copenhagen, Denmark; Dept. of Surgical Oncology, Princess Margaret Cancer Centre, Toronto ON M5G 2M9, Canada; Dept. of Surgery (Urology), University of Toronto, Canada; Institute of Environmental Medicine, Karolinska Institutet, 177 77 Stockholm, Sweden; Division of Cancer Epidemiology and Genetics, National Cancer Institute, NIH, Bethesda, Maryland, 20892, USA; Cancer Epidemiology Unit, Nuffield Department of Population Health, University of Oxford, Oxford, UK; Division of Epidemiology, Department of Medicine, Vanderbilt University Medical Center, 2525 West End Avenue, Suite 800, Nashville, TN 37232 USA; International Epidemiology Institute, Rockville, MD 20850, USA; Department of Epidemiology, Harvard T. H. Chan School of Public Health, Boston, MA 02115, USA; Sorbonne Universite, GRC n°5, AP-HP, Tenon Hospital, 4 rue de la Chine, F-75020 Paris, France; CeRePP, Tenon Hospital, F-75020 Paris, France; Department of Molecular Medicine, Aarhus University Hospital, Palle Juul-Jensen Boulevard 99, 8200 Aarhus N, Denmark; Department of Clinical Medicine, Aarhus University, DK-8200 Aarhus N; International Hereditary Cancer Center, Department of Genetics and Pathology, Pomeranian Medical University, 70-115 Szczecin, Poland; Exposome and Heredity, CESP (UMR 1018), Faculté de Médecine, Université Paris-Saclay, Inserm, Gustave Roussy, Villejuif; Department of Cancer Epidemiology, Moffitt Cancer Center, 12902 Magnolia Drive, Tampa, FL 33612, USA; Cancer Epidemiology Division, Cancer Council Victoria, 200 Victoria Parade, East Melbourne, VIC, 3002, Australia; Centre for Epidemiology and Biostatistics, Melbourne School of Population and Global Health, The University of Melbourne, Grattan Street, Parkville, VIC 3010, Australia; Department of Radiation Oncology and Department of Genetics and Genomic Sciences, Box 1236, Icahn School of Medicine at Mount Sinai, One Gustave L. Levy Place, New York, NY 10029, USA; Centre for Cancer Biomarker and Biotherapeutics, Barts Cancer Institute, Queen Mary University of London, John Vane Science Centre, Charterhouse Square, London, EC1M 6BQ, UK; Uro Care, Kampala, Uganda; Fundación Pública Galega Medicina Xenómica, Santiago de Compostela, 15706, Spain; Instituto de Investigación Sanitaria de Santiago de Compostela, Santiago de Compostela, 15706, Spain; Centro de Investigación en Red de Enfermedades Raras (CIBERER), Spain; Lineberger Comprehensive Cancer Center, University of North Carolina at Chapel Hill, 450 West Drive, CB 7295, Chapel Hill, NC 27599, USA; Department of Epidemiology, University of North Carolina at Chapel Hill, Chapel Hill, NC, USA; Epidemiology Branch, National Institute of Environmental Health Sciences, Research Triangle Park, NC, USA; http://impact.icr.ac.uk; ISGlobal, Barcelona, Spain; IMIM (Hospital del Mar Medical Research Institute), Barcelona, Spain; Universitat Pompeu Fabra (UPF), Barcelona, Spain; CIBER Epidemiología y Salud Pública (CIBERESP), 28029 Madrid, Spain; Department of Medical Epidemiology and Biostatistics, Karolinska Institutet, SE-171 77 Stockholm, Sweden; Department of Laboratory Genetics, Portuguese Oncology Institute of Porto (IPO Porto) / Porto Comprehensive Cancer Center, Porto, Portugal; Cancer Genetics Group, IPO Porto Research Center (CI-IPOP) / RISE@CI-IPOP (Health Research Network), Portuguese Oncology Institute of Porto (IPO Porto) / Porto Comprehensive Cancer Center, Porto, Portugal; School of Medicine and Biomedical Sciences (ICBAS), University of Porto, Porto, Portugal; Univ Rennes, Inserm, EHESP, Irset (Institut de recherche en santé, environnement et travail) - UMR_S 1085, Rennes, France; Department of Cell Systems and Anatomy, Mays Cancer Center, University of Texas Health Science Center at San Antonio, San Antonio Texas; Cancer Prevention Graduate School, German Cancer Research Center (DKFZ), D-69120, Heidelberg, Germany; Network Aging Research (NAR), Heidelberg University, D-69120 Heidelberg, Germany; Departments of Epidemiology & Population Health and of Medicine, Division of Oncology, Stanford Cancer Institute, Stanford University School of Medicine, Stanford, CA 94304 USA; Molecular Medicine Center, Department of Medical Chemistry and Biochemistry, Medical University of Sofia, Sofia, 2 Zdrave Str., 1431 Sofia, Bulgaria; The University of Texas M. D. Anderson Cancer Center, Department of Genitourinary Medical Oncology, 1515 Holcombe Blvd., Houston, TX 77030, USA; Department of Population Sciences, Beckman Research Institute of the City of Hope, 1500 East Duarte Road, Duarte, CA 91010; Department of human structure and repair, Ghent University, Ghent, Belgium; Department of Surgery, Faculty of Medicine, University of Malaya, 50603 Kuala Lumpur, Malaysia; Division of Epidemiology, Department of Preventive Medicine, University of Tennessee Health Science Center, Memphis, TN 38163; Department of Oncology, University Hospital Centre Zagreb, University of Zagreb, School of Medicine, 10 000 Zagreb, Croatia; Department of Oncology, Cross Cancer Institute, University of Alberta, 11560 University Avenue, Edmonton, Alberta, Canada T6G 1Z2; Division of Radiation Oncology, Cross Cancer Institute, 11560 University Avenue, Edmonton, Alberta, Canada T6G 1Z2; Molecular Endocrinology Laboratory, Department of Cellular and Molecular Medicine, KU Leuven, BE-3000, Belgium; Genetic Oncology Unit, CHUVI Hospital, Complexo Hospitalario Universitario de Vigo, Instituto de Investigación Biomédica Galicia Sur (IISGS), 36204, Vigo (Pontevedra), Spain; Uniformed Services University, 4301 Jones Bridge Rd, Bethesda, MD 20814, USA; Center for Prostate Disease Research, 6720A Rockledge Drive, Suite 300, Bethesda, MD 20817, USA; Epidemiology and Biostatistics Unit, Centre Armand-Frappier Santé Biotechnologie, Institut national de la recherche scientifique, 531 Boul. des Prairies, Laval, QC, Canada H7V 1B7; Department of Social and Preventive Medicine, School of Public Health, University of Montreal, Montreal, QC, Canada; The University of Miami School of Medicine, Sylvester Comprehensive Cancer Center, 1120 NW 14th Street, CRB 1511, Miami, Florida 33136, USA; http://www.cancerresearchuk.org/about-cancer/find-a-clinical-trial/a-study-find-out-looking-gene-changes-would-be-useful-in-screening-for-prostate-cancer-profile-pilot; http://www.icr.ac.uk/our-research/research-divisions/division-of-genetics-and-epidemiology/oncogenetics/research-projects/ukgpcs/ukgpcs-collaborators; The Institute of Cancer Research, London, SM2 5NG, UK; Royal Marsden NHS Foundation Trust, London, SW3 6JJ, UK; Corporal Michael Crescenz Veterans Affairs Medical Center, Philadelphia, PA, USA; Department of Medicine - Hematology/Oncology, Abramson Cancer Center, University of Pennsylvania, Philadelphia, PA, USA; Division of General Internal Medicine and Primary Care, VA Boston Healthcare System, Boston, MA, USA; Departments of Radiology, University of California San Diego, La Jolla, CA, USA; Department of Bioengineering, University of California San Diego, La Jolla, CA, USA; Department of Urology, University of California San Diego, La Jolla, CA, USA

**Keywords:** prostate cancer, polygenic score, germline, genomics, active surveillance, Gleason score, GWAS

## Abstract

**Background:** Risk stratification for prostate cancer (PCa) progression or aggressiveness is often based on clinicopathologic features, some of which may be influenced by genetic factors. We developed a novel, germline polygenic risk score (PRSagg) to predict likelihood of developing aggressive PCa.

**Methods:** PRSagg was developed using data from 38,688 patients with PCa (case-only analysis) from the Million Veteran Program (MVP) through a genome-wide search for variants associated with PCa grade group at diagnosis. We tested associations of PRSagg with grade group using the entire MVP dataset using the .632 bootstrap method. In an MVP cohort with localized PCa that was initially monitored without treatment, we tested PRSagg for association with unfavorable outcomes (subsequent development of grade group 4-5, metastasis, and/or biochemical recurrence after definitive treatment). We performed external validation in data from patients in the PRACTICAL Consortium (n=45,214) and from participants in the ProtecT randomized trial who underwent active monitoring (n=316). Odds ratios (ORs) were calculated per standard deviation (SD) increase with 95% confidence intervals, while adjusting for age, genetic ancestry, a previously developed polygenic score for risk of PCa (PHS601), and a polygenic score for benign elevated prostate-specific antigen (PRS_PSA_). For the outcome of metastasis, we additionally adjusted for PSA at diagnosis.

**Results:** In the MVP training dataset, PRSagg (172 variants) was associated with higher grade group at diagnosis (OR = 1.53 [1.51-1.56]) and with increased risk of unfavorable outcomes during monitoring (OR = 1.13 [1.09-1.18]). These findings were confirmed in the external datasets. PRSagg was associated with greater odds of higher grade group at diagnosis (OR = 1.09 [1.06-1.11]). Among ProtecT participants undergoing active monitoring, PRSagg was associated with higher risk of metastasis (OR = 2.15 [1.02-3.88]). Among MVP participants with high polygenic risk of developing any PCa, the risk of aggressive disease was highest in men with high PRSagg and low genetic risk of PSA elevation.

**Conclusions:** Among men who develop PCa, a weighted sum of common germline variants (PRSagg) is independently associated with PCa aggressiveness. These findings may inform future study of germline influence on tumor evolution and risk-stratified intensity of active surveillance.

## Introduction

Prostate cancer (PCa) is common and a major public health challenge^1^. While many cases of PCa are indolent and unlikely to cause significant problems in a patient’s lifespan, some patients develop aggressive disease, and PCa remains a leading cause of cancer death^1,2^. Patients with lower-grade localized PCa often undergo active surveillance (AS), avoiding the side effects of immediate treatment while still keeping the window open for curative treatment^3^. However, some patients experience “AS failure,” which has been defined as pathological upgrade, metastasis, or recurrent cancer after treatment^4^. It is unclear how to best risk-stratify patients diagnosed with lower-grade PCa and to what extent inherited factors might be associated with aggressiveness of PCa.

Germline genotyping and sequencing are increasingly available, affording new risk stratification opportunities^5^. Some rare pathogenic germline variants are associated with an elevated lifetime risk of developing PCa, and a few (e.g., *BRCA2*) are associated with developing more aggressive PCa^6^. Meanwhile, many common genetic variants have a small effect size individually, but combinations of multiple common variants can yield polygenic scores that achieve meaningful risk stratification. A number of studies using genetic risk scores have already demonstrated clinically meaningful risk stratification for development of PCa^7–14^. Unlike family history and self-reported race, genetic scores offer a more objective, quantitative way to assess risk. In addition, germline genetic scores, unlike prostate-specific antigen (PSA) or MRI, only need to be calculated once in a patient’s lifetime. Recently, our group has used genotype and phenotype data from the Million Veteran Program (MVP) to develop PHS601, a polygenic hazard score associated with earlier age at diagnosis of PCa. This score is being used in an ongoing national randomized controlled trial of genomics-informed, precision PCa screening (ProGRESS; NCT05926102)^15^.

Polygenic scores like PHS601 are associated with development of any PCa, both indolent and aggressive^10,12^. A pertinent clinical and scientific question is whether germline genetic variants also influence PCa aggressiveness^16^. For example, a polygenic score associated with cancer aggressivity might provide useful information in the setting of AS^17^. Early detection methods could benefit patients by anticipating the most severe PCa outcomes (thus reducing the risk of overdiagnosis). Challenges to creating a score specific for aggressive PCa may be partially due to data and methodological limitations. For example, case-control datasets used to develop polygenic scores often lacked detailed information on the aggressiveness of disease at diagnosis or long-term follow-up^5^. Another potential limitation is the common practice of selecting arbitrary criteria (e.g., grade group ≥2)) to define non-aggressive vs. aggressive cancers, whereas, in clinical practice, cancers are graded along a broader spectrum. Also, low-risk cancers have historically often been treated radically, making it impossible to know which of these would have developed high-grade cancer later in their lifetime. Nevertheless, there is evidence that polygenic scores could provide independent value in predicting benign elevations in serum PSA and patient PCa outcomes^16–18^.

Leveraging data from a large, longitudinal cohort with linkage to detailed clinical records, we developed a new polygenic risk score, called PRSagg, designed to be predictive of PCa grade group—independent of age at diagnosis, genetic ancestry, polygenic risk of any PCa (PHS601), and a polygenic score for PSA (PRS_PSA_). Grade group is widely understood to be one of the most important indicators of PCa aggressiveness and drives key treatment decisions^19,20^. We evaluated whether men with higher PRSagg are at increased risk of higher grade group. We also tested whether PRSagg is associated with risk of AS failure (defined as subsequent development of grade group 4-5, metastasis, or biochemical recurrence after treatment with surgery or radiotherapy)^4^. Findings were validated in two independent datasets. This work could yield insights into the potential for polygenic scores to inform tailored AS intensity and the relationship of inherited risk with PCa aggressiveness.

## Methods

### Million Veteran Program

Data from the Million Veteran Program (MVP) were used to develop the polygenic risk score for aggressive prostate cancer (PRSagg) using a case-only design. We identified 38,688 MVP male participants with a diagnosis of PCa and a Gleason score obtained via biopsy within 1 year of the date of diagnosis. Clinical data–including age at diagnosis, Gleason score, and date of Gleason score–were retrieved from the VA Corporate Data Warehouse based on International Classification of Diseases (ICD) diagnosis codes and VA Central Cancer Registry data^14^. Details on MVP data collection, genotyping, genetic imputation, and quality control have been described previously^14^. Dosages for all variants used in this study were extracted from imputed data using the TOPMed reference panel.

### Candidate genetic variants

First, we conducted a genome-wide SNP search for candidate variants associated with grade group, the primary outcome of interest for PRSagg. Initially, SNPs were pruned to those with a minimum minor allele frequency of 0.05, as well as those in linkage equilibrium with each other using a window size of 100 kb and R^2^ threshold of 0.8. Next, a general linear model was fit at each variant using the grade group (coded as a numeric variable from 1 to 5) as the phenotype/outcome variable, and a covariate matrix including: age at PCa diagnosis, a polygenic hazard score for PCa (PHS601)^15^, a polygenic risk score for benign elevated PSA (PRS_PSA_)^21^, and the first 10 principal components of genetic ancestry estimated from the FastPop ancestry informative markers^12,13,22^. SNPs with adjusted *p*-values <10^-4^ were selected for further analysis.

In parallel, we conducted a case-case GWAS to contribute to a larger consortium meta-analysis aimed at identifying novel variants associated with aggressive PCa. This GWAS used methods agreed on in the larger consortium and differ somewhat from our SNP search. Aggressive PCa was defined as meeting one or more of the following criteria: Gleason grade group 4 or 5 PCa at diagnosis; PSA at diagnosis of >20ng/ml; N1 stage; or M1 stage. Non-aggressive cases were defined as individuals with all of the following at diagnosis: Gleason grade group 1 or 2 PCa; PSA <10ng/ml; N0 and M0 stage disease. Intermediate cancers (grade group 3 or grade group 1-2 with PSA 10-20 ng/mL) were excluded from this analysis to more starkly compare the extremes. The GWAS was performed using MVP Release 4 TopMed data (GRCh38). Principal components were calculated using plink2 on genotyped data, and association testing was performed with SAIGE v1.3.0 using age and the first 10 principal components as covariates. SNPs from this GWAS with *p*-values <10^-6^ were added to the initial SNP search results (after removing any duplicates) to form the final list of candidate variants.

### Model fitting

A LASSO-regularized general linear model was fit using the grade group (coded as a numeric variable from 1 to 5; ordinal outcome variables are not supported in the *GLMNET* package of R) as the outcome variable and a predictor matrix including: dosages (0,1, 2) for the search SNPs as the primary exposure, along with the following covariates: polygenic risk of any PCa (PHS601), PRS_PSA_, age at PCa diagnosis, and genetic ancestry grouping (European, African, East Asian, and Admixed American). Regularization was restricted to the search SNP dosages. The final form of the LASSO model was estimated using the lambda value that was within 1 standard error of the minimum mean cross-validated error. PRSagg was estimated for each patient as the matrix multiplication product of the search SNP dosages and the corresponding coefficients derived from the LASSO model.

### Testing PRSagg in MVP

PRSagg was first evaluated in MVP (the training dataset) in two scenarios: (1) association with PCa grade group using the entire training dataset; and (2) association with unfavorable outcomes (similar to AS failure defined previously^4^) in a cohort with localized PCa that did not undergo immediate treatment.

#### Association with grade group

A proportional odds logistic regression model was fit using the grade group as an ordered categorical variable and predictors including PRSagg, PHS601, PRS_PSA_, age at PCa diagnosis, and genetic ancestry grouping. The generalization performance of the model was estimated using the .632 bootstrap method, which has been shown to outperform cross-validation^23^. Briefly, for each of 100 bootstraps, a training and out-of-bag sample were generated. The PRSagg model was fit, as described above using the LASSO model, in the bootstrap training set and tested in the out-of-bag sample using the proportional odds logistic regression model. The coefficients for the predictor variables were pulled for each of the 100 out-of-bag samples. The final validation performance coefficients were then estimated as the weighted sum of the out-of-bag coefficient (weight: 0.632) and full dataset coefficient (weight: 0.368). Point-estimates of the mean and corresponding 95% confidence intervals were generated from the bootstrapped distribution^24^.

#### Association with unfavorable outcomes during monitoring

A cohort approximating AS or active monitoring was identified from all MVP male participants (MVP Monitored PCa Cohort) and was defined as those individuals: (1) with a known PCa diagnosis; (2) whose first recorded treatment (prostatectomy, radiation therapy, or androgen deprivation therapy) was more than one year after diagnosis; and (3) whose first recorded grade group was 1 or 2. In total, 10,257 men met these criteria. An unfavorable outcome was classified according to a previously published definition of “active surveillance failure,” which includes any one of: subsequent diagnosis with Gleason grade group 4 or 5, diagnosis of metastatic PCa, or rising PSA following treatment (ICD-10 code R97.21)^4^. A generalized linear model with logit link function was fit with unfavorable outcomes as the binary outcome variable, and a predictor matrix including PRSagg, PHS601, PRS_PSA_, age at PCa diagnosis, and genetic ancestry grouping. Patients who had not experienced the event by last follow-up or death from other cause were considered to not have experienced the event for this logistic regression analysis. As the primary objective was to explore whether there is any association between germline polygenic markers and PCa aggressiveness, we focused on odds ratios in multivariable models. The datasets available are likely not adequate for robust conclusions regarding added clinical utility for risk stratification; nonetheless, area under the curve (AUC) analyses are described in the supplementary material. Calibration curves and decision analysis are deferred to larger datasets necessary for robust evaluation of clinical utility. In addition, the above linear model was fit with log10-transformed PSA as an additional predictor, in the subset of individuals with an available PSA in the year before diagnosis. Confidence intervals of 95% were estimated using a standard bootstrapping of the dataset in this and all subsequent modeling analyses.

### PRACTICAL dataset

Genetic and phenotype data from the PRACTICAL Consortium were used as an external validation dataset for PRSagg. In total, we identified 44,802 men with a diagnosis of PCa and an available Gleason score. As primary and secondary Gleason patterns were not available for most of the PRACTICAL dataset, grade groups were collapsed into four possible values for grade group: 1, 2-3, 4, or 5. Dosages for all variants used in this study were extracted from imputed data using the 1000 Genomes Phase 3 reference panel.

#### Association with PCa grade group

The association between PRSagg and grade group at diagnosis, coded as an ordered categorical variable, was estimated using a proportional odds logistic regression model, as before, while including the following as covariates: PHS601, PRS_PSA_, age at prostate cancer diagnosis and genetic ancestry grouping (European, African, East Asian).

### ProtecT active monitoring dataset

#### Association with unfavorable outcomes during AM

We identified a cohort of participants in the ProtecT randomized controlled trial who underwent AM and had available genetic data (n = 316)^2,7,25^. The primary outcome of interest was metastasis. As metastasis was a rare event^2^, we also secondarily evaluated progression to stage ≥T3 and transition out of AM to check for consistency in direction of effect. For each outcome, a generalized linear model was fit using a predictor matrix including PRSagg, PHS601, PRS_PSA_, age at PCa diagnosis, and PSA at randomization.

### Pathway analysis

We performed a pathway analysis for the set of identified genes that contain or are proximal to our PRSagg set of variants using the STRING database^26^.

### Ensemble analysis

Bioinformatics analyses was performed using Ensembl (USC Genome Browser)^27^. SNP annotations results including PCa risk contribution were generated.

## Results

### Patient characteristics for MVP datasets

Patient characteristics for the MVP datasets are shown in Table 1.

**Table 1.** Breakdown of the MVP datasets used in training and testing the model. Values for age, PSA, and years of follow-up are given as median [interquartile range]; year of diagnosis is given as range (min-max); the other rows are listed as number of individuals (% of total N in this column). Eligibility for the Training dataset were: grade group and age both available within one year of first diagnosis. Eligibility for the MVP Monitored PCa Cohort were: no treatment in the first year of diagnosis with PCa that was not grade group 3-5 at diagnosis. There were 5,776 patients who met eligibility criteria for both cohorts.

|  | Training | MVP Monitored PCa Cohort |
| --- | --- | --- |
| N | 38,688 | 10,257 |
| Age at diagnosis (years) | 66.4 [61.6 – 71.3] | 65 [60 – 69.8] |
| Year of diagnosis | 1998-2024 | 1965-2023 |
| PSA at diagnosis (ng/mL) | 6.27 [4.71 – 9.43] | 5.33 [4.1 – 7.33] |
| Years of follow-up | 8.9 [5.1 - 13.5] | 12.4 [8.5-17.0] |
| Genetic Ancestry |  |  |
| EUR | 24,205 (62.5%) | 6,807 (66.4%) |
| AFR | 11,836 (30.6%) | 2,822 (27.5 %) |
| AMR | 2,458 (6.35%) | 578 (5.64%) |
| EAS | 189 (0.48%) | 50 (0.49 %) |
| Grade Group<sup>1</sup> |  |  |
| 1 | 15,095 (39.0%) | 5,944 (58.0%) |
| 2 | 10,842 (28.0%) | 2,693 (26.3%) |
| 3 | 5,024 (12.9%) | 548 (5.34%) |
| 4 | 3,980 (10.3%) | 567 (5.52%) |
| 5 | 3,747 (9.69%) | 505 (4.92%) |

<sup>1</sup> First-available grade group. The training set was limited to those individuals whose first available grade group was within the first year after diagnosis.

### Candidate genetic variants

The SNP search identified 186 candidate variants associated with grade group. The case-case GWAS identified 16 variants associated with aggressive PCa (by comparing 1,132 cases of aggressive PCa to 27,645 cases of non-aggressive PCa in MVP, using the aggressive PCa consortium definitions). 4 of the 172 PRSagg variants overlapped with variants in PHS601.

The 172 variants that were selected for inclusion in the final version of PRSagg are listed in Supplementary Table 1. For each variant, we list the chromosome, position (in GrCh38 and GRCh37 builds), effect and reference alleles, PRSagg model coefficients, and effect allele frequencies (for the entire training set, men of European genetic ancestry, and men of African genetic ancestry).

### Performance in MVP

PRSagg was associated with higher grade group at diagnosis in the training set, when adjusting for age, genetic ancestry, PHS601, and PRS_PSA_ (see Table 2 for coefficients). For every standard deviation increase in PRSagg, the odds of being in a higher grade group increased by 53%. Additional analyses suggest the proportional odds assumption was reasonable (Supplementary Material).

**Table 2.** Odds ratios (and 95% confidence intervals) of the variables used in testing the association between PRSagg and grade group in the MVP training dataset, using a proportional odds logistic regression model. OR for polygenic scores are in units of SD.

| Variable | OR [95%CI] |
| --- | --- |
| PRSagg | 1.53 [1.51-1.56] |
| Age at diagnosis (years) | 1.06 [1.06-1.06] |
| PHS601 | 1.05 [1.03 - 1.08] |
| PRS<sub>PSA</sub> | 0.87 [0.85 - 0.92] |
| Genetic Ancestry |  |
| AFR | 1.09 [1.05 - 1.14] |
| AMR | 1.19 [1.11 - 1.26] |
| EAS | 1.30 [0.97 - 1.64] |

PRSagg also was modestly associated with unfavorable outcomes in the MVP Monitored PCa Cohort, while accounting for age at diagnosis, genetic ancestry, PHS601, and PRS_PSA_ (see Table 3). For every standard deviation increase in PRSagg, the odds of an unfavorable outcome increased by 13% (defined as subsequent development of Gleason grade group 4 or 5 on subsequent biopsy during monitoring, diagnosis of metastatic PCa, or biochemical recurrence following subsequent definitive treatment). PRSagg remained significantly associated with unfavorable outcomes after including the log-10-transformed PSA at diagnosis (n = 6,898). (Supplementary Table 2).

**Table 3.** Odds ratios (and 95% confidence intervals) of the variables used in testing the association between PRSagg and unfavorable outcomes in the MVP Monitored PCa Cohort, using a generalized linear model with a logit link function. OR for polygenic scores are in units of SD.

| Variable | OR [95%CI] |
| --- | --- |
| PRSagg | 1.13 [1.09-1.18] |
| Age at diagnosis (years) | 0.99 [0.99-1.00] |
| PHS601 | 1.03 [0.97-1.07] |
| PRS<sub>PSA</sub> | 0.94 [0.91-0.98] |
| Genetic Ancestry |  |
| AFR | 0.85 [0.77-0.96] |
| AMR | 1.09 [0.89-1.31] |
| EAS | 1.12 [0.63-1.95] |

### Patient characteristics for external validation datasets

Patient characteristics of PRACTICAL and ProtecT active monitoring datasets are tabulated in Table 4 and Table 5, respectively.

**Table 4.** Patient characteristics of PRACTICAL dataset used as an independent validation set for the association between PRSagg and grade group. Values for age and PSA are given as median [interquartile range]; the other rows are listed as number of individuals (% of total N in this column).

|  | PRACTICAL |
| --- | --- |
| N | 44,802 |
| Age at diagnosis (years) | 64.6 [59.0 – 69.9] |
| PSA at diagnosis (ng/mL) | 7.1 [4.7-12.7] |
| Genetic Ancestry |  |
| EUR | 41,052 (91.6%) |
| AFR | 2,917 (6.51%) |
| EAS | 833 (1.86%) |
| Grade Group<sup>1</sup> |  |
| 1 | 20,678 (46.2%) |
| 2/3 | 17,397 (38.8%) |
| 4 | 3,459 (7.72%) |
| 5 | 3,268 (7.29%) |

<sup>1</sup> First-available grade group. The training set was limited to those individuals whose first available grade group was within the first year after diagnosis.

**Table 5.** Patient characteristics of ProtecT active monitoring (AM) dataset used as an independent validation set for the association between PRSagg and unfavorable outcomes while on AM. Age, PSA, and Gleason score are those at randomization. Values for age, PSA, and years of follow-up are given as median [interquartile range]; the other rows are listed as number of individuals (% of total N in this column).

|  | Protect AM |
| --- | --- |
| N | 316 |
| Age at diagnosis (years) | 63.9 [59.1-67.4] |
| PSA (ng/mL) | 4.5 [3.6-6.4] |
| Years of follow-up | 4.7 [0.0-9.0] |
| Genetic Ancestry |  |
| EUR | 316 (100%) |
| Gleason Score |  |
| 2+2 | 1 (0.32%) |
| 3+3 | 250 (79.1%) |
| 3+4 | 49 (15.5%) |
| 4+3 | 11 (3.48%) |
| 3+5 | 1 (0.32%) |
| 4+4 | 3 (0.95%) |
| 4+5 | 1 (0.32%) |
| Outcomes |  |
| Metastases | 27 (8.54%) |
| Progression to ≥ T3 | 60 (19.0%) |
| Transition out of AM | 136 (43.0%) |

### Performance in external validation datasets

PRSagg was independently associated with higher grade group at diagnosis in PRACTICAL (Table 6), after accounting for standard clinicopathologic variables. For every standard deviation increase in PRSagg, the odds of being in a higher grade group increased by 9%. In ProtecT, PRSagg was independently associated with each of the unfavorable AM outcomes: metastasis, progression to stage >= T3, and transition out of AM to PCa treatment (Table 7, Supplementary Table 3).

**Table 6.** Odds ratios (and 95% confidence intervals) of the variables used in testing the association between PRSagg and grade group in PRACTICAL, using a proportional odds logistic regression model. OR for polygenic scores are in units of SD.

| Variable | OR [95% CI] |
| --- | --- |
| PRSagg | 1.09 [1.06-1.11] |
| Age at diagnosis (years) | 1.02 [1.01-1.02] |
| PHS601 | 1.00 [0.98 - 1.02] |
| PRS<sub>PSA</sub> | 0.88 [0.87 - 0.90] |
| Genetic Ancestry |  |
| AFR | 1.09 [1.02 - 1.17] |
| EAS | 1.46 [1.29 - 1.67] |

**Table 7.** Odds ratios (and 95% confidence intervals) of the variables used in testing the association between PRSagg and metastasis in ProtecT, using a generalized linear model with a logit link function. OR for polygenic scores are in units of SD.

| Variable | OR [95% CI] for Metastasis? |
| --- | --- |
| PRSagg | 2.15 [1.02-3.88] |
| Age at diagnosis (years) | 1.03 [0.91-1.16] |
| PSA | 1.17 [1.02-1.37] |
| PHS601 | 1.24 [0.72-1.90] |
| PRS<sub>PSA</sub> | 0.56 [0.32-0.87] |

### PRSagg/psa groups

Genetic aggressivity risk groups were made by combining the two polygenic scores that were consistently found to be associated with higher risk of aggressive disease across datasets: PRS_PSA_ and PRSagg (as PRSagg was developed with PRS_PSA_ as a covariate, the two scores are not correlated; *R^2^* = 0.001). As an illustration, each score was first subset into three risk categories: low (bottom quartile), mid (25th - 75th percentile), and high (top 25th percentile). Figure 1 shows the cumulative incidence of a grade group ≥ 3 at diagnosis in the MVP training set for every risk category combination. Genetic aggressivity risk groups (PRSagg/psa groups) were defined as:

- low if PRSagg is low and PRS_PSA_ is high
- mid if PRSagg is mid and PRS_PSA_ is mid
- high if PRSagg is high and PRS_PSA_ is low

**Figure 1.**
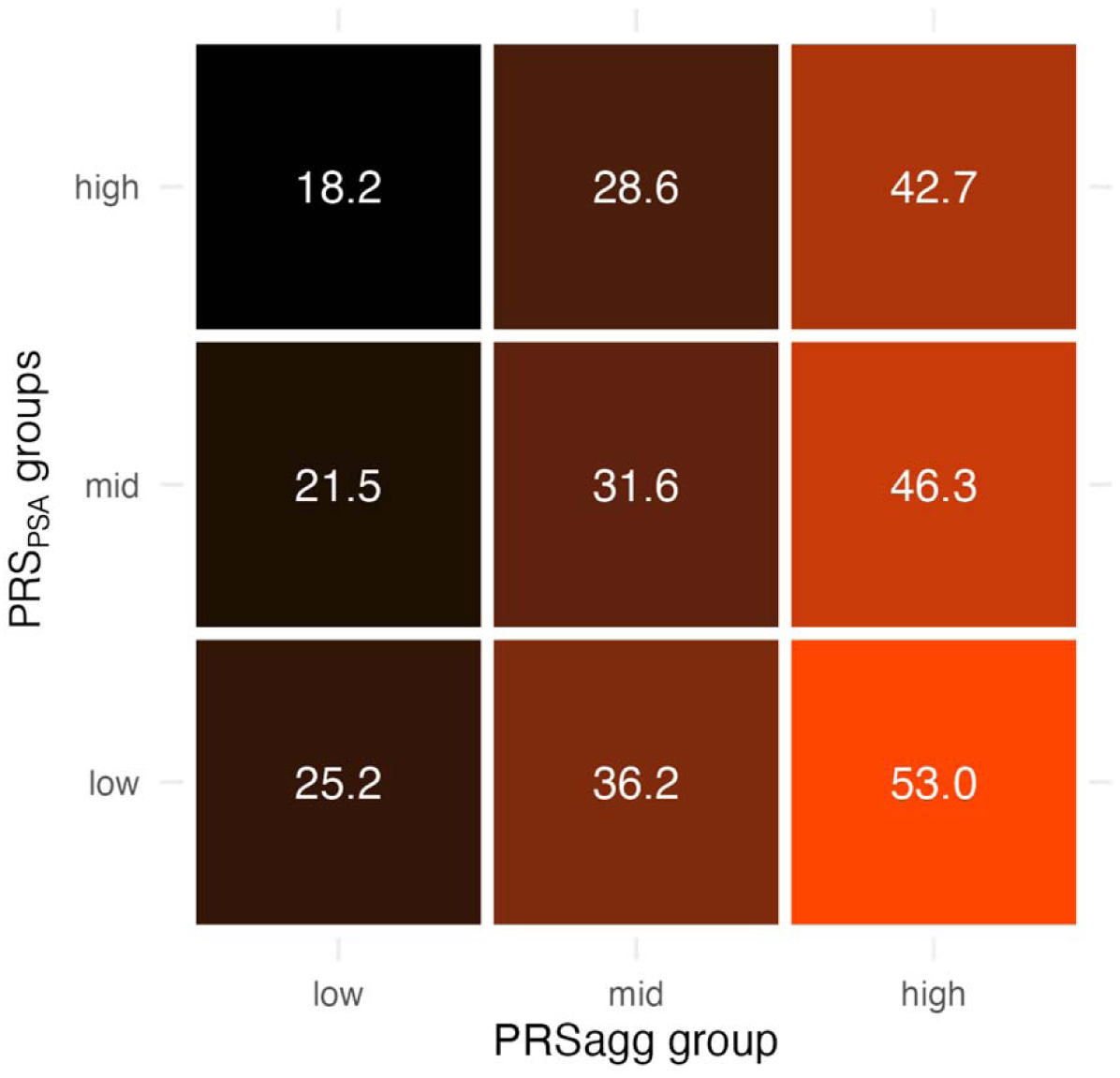
Cumulative incidence (%) of grade group >=3 for every combination of PRSagg and PRS_PSA_ risk category in the MVP training dataset.

Using this definition of PRSagg/psa group, we fit a Cox proportional hazards model in MVP data using the age at diagnosis of metastatic cancer as the time-to-event outcome and a predictor matrix including genetic ancestry, PRSagg/psa group, and PHS601 risk categories (as previously defined). Individuals without metastatic cancer were censored at age of last follow-up or non-PCa death. The cumulative incidence of metastatic prostate cancer by age 85 years for men of European genetic ancestry, and various combinations of PRSagg/psa group and PHS601 risk categories are shown in Figure 2. In Figure 3, the entire cumulative incidence curve between ages 45 and 90 years for men in the high PHS601 risk category across genetic ancestry (European, African) and PRSagg/psa group (low, mid, high) are plotted.

**Figure 2.**
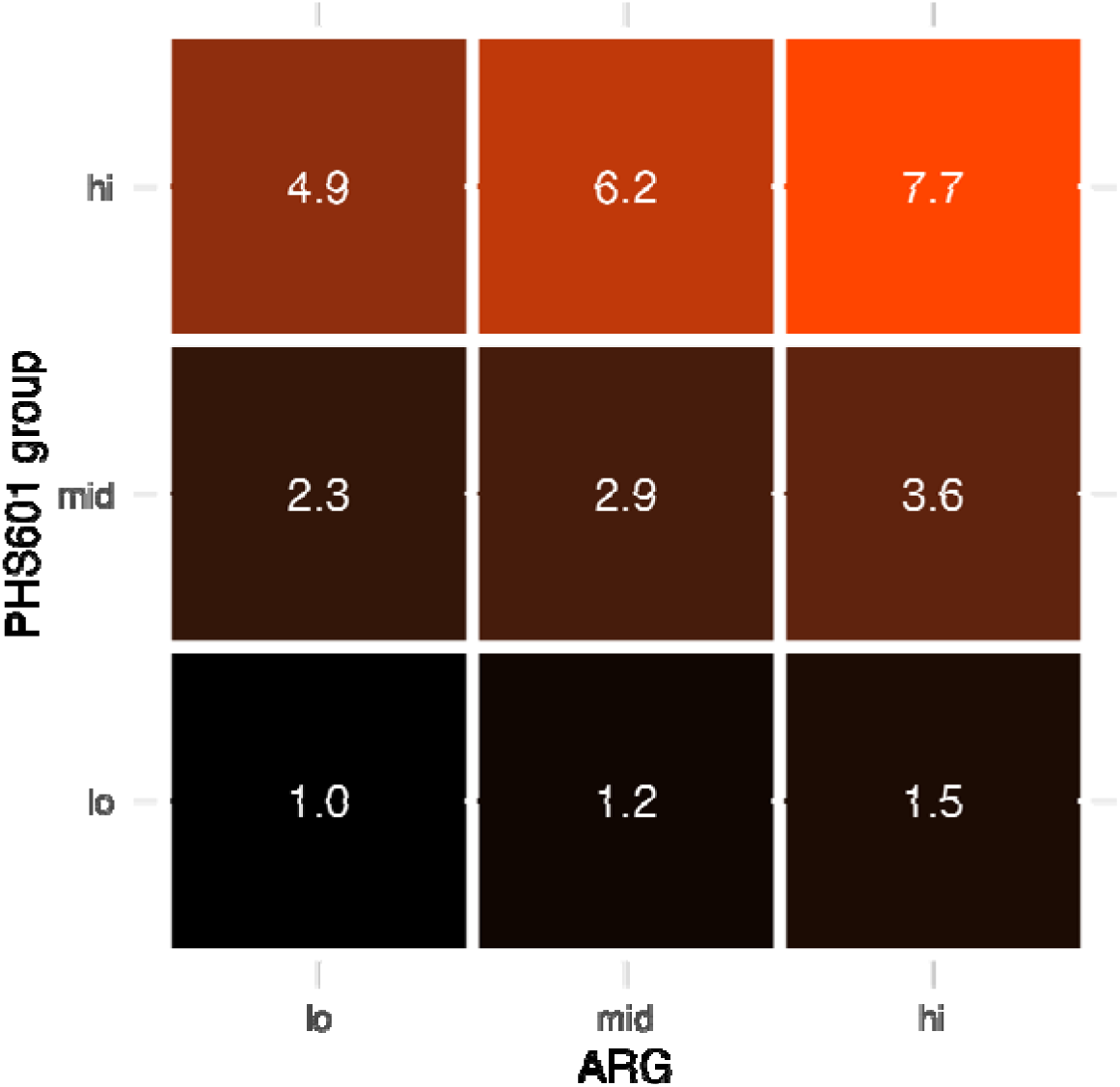
Cumulative incidence of metastatic prostate cancer by age 85 years for men of European genetic ancestry in MVP, across PRSagg/psa group and PHS601 risk groups in the full MVP dataset.

**Figure 3.**
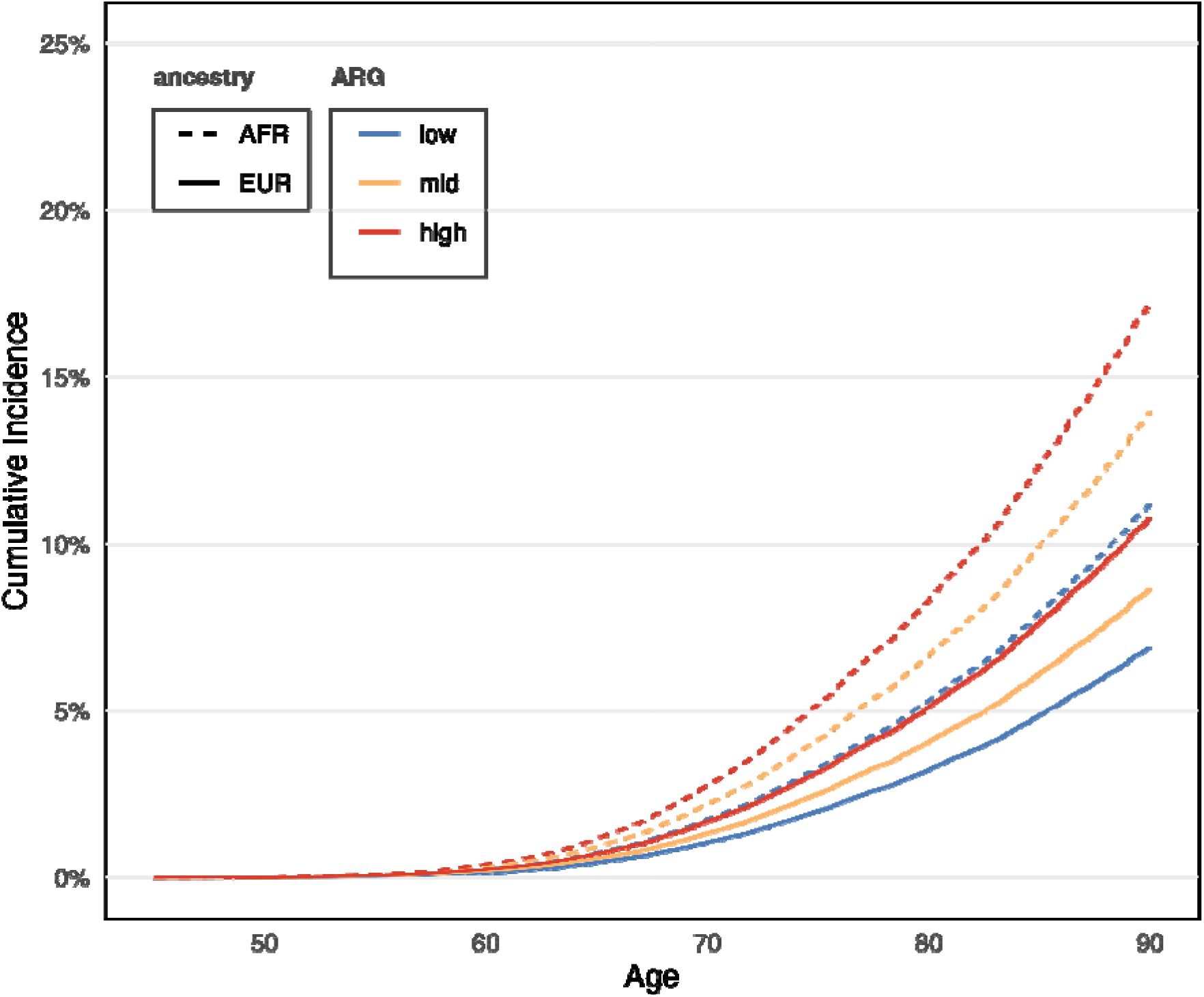
Cumulative incidence of metastatic PCa among MVP participants who have a high genetic risk of any PCa (i.e., high PHS601), across genetic ancestry (AFR: African, EUR: European) and genetic aggressivity risk group (PRSagg/psa group, a combination of polygenic risk of aggressive prostate cancer, PRSagg, and polygenic risk of elevated PSA, PRS_PSA_). Genetic ARG further stratifies risk of metastasis beyond genetic ancestry and PHS601.

### STRING pathway analysis

Interactions identified by the pathway analysis from the STRING database appear in Supplementary Table 6 and are graphically depicted in Supplementary Figure 1.

### Ensembl Gene analysis

SNP annotations results, including genes, biological function, and PCa aggressive risk contribution, are described in Supplementary Table 7.

## Discussion

We used common germline markers to develop a polygenic score (PRSagg) associated with PCa aggressiveness. This score was independently associated with increasing grade group when accounting for age, genetic ancestry, polygenic risk of any PCa, and polygenic predisposition to benign PSA elevation. As grade group is strongly associated with risk of metastasis, common genetic variation present at birth is related to prognosis among those who develop PCa. Beyond the potential biological implications of this finding, there may be clinical value in polygenic scores at time of diagnosis with grade group 1 or 2 PCa, which often does not require immediate treatment^4,28^. Among participants of the ProtecT randomized controlled trial assigned to active monitoring, we observed a 2.15-fold increased risk of developing metastasis per standard deviation of PRSagg. We also observed a 44% reduction of risk per standard deviation of PRS_PSA_. Effects in the same direction, albeit smaller magnitude, were found in a cohort of monitored PCa cases identified retrospectively within MVP. All these findings were from multivariable models accounting for common clinical variables. Taken together, this study makes an intriguing case for investigation into potential mechanistic underpinnings of germline association with PCa aggressiveness. Likewise, prospective studies aiming to optimize active surveillance could incorporate polygenic risk scores.

As both PRSagg and PRS_psa_ were associated with PCa aggressiveness, we illustrated their combined effects by forming broad categories (PRSagg/psa group) based on low, middle or high quartiles of each polygenic score. Among all MVP participants, the absolute risk of grade group ≥3 PCa by age 85 years ranged from 18.2% for those with low PRSagg/psa group (low PRSagg and high PRS_PSA_) to 53.0% for those with high PRSagg/psa group (high PRSagg and low PRS_PSA_). Combining genetic risk of any PCa (PHS601) with genetic risk of aggressive PCa (ARG) stratified overall risk of developing metastatic PCa. Among European-ancestry MVP participants, those with low PHS601 and low PRSagg/psa group had a cumulative incidence of metastatic PCa of 1.0% by age 85 years, while those with high PHS601 and high PRSagg/psa group had a cumulative incidence of 7.7%, a nearly 8-fold increase. Thus, PRSagg and PRS_PSA_ might add meaningful value in predicting lifetime risk of metastatic PCa beyond the already powerful prediction possible for any PCa achieved with scores like PHS601, Larger datasets should permit evaluation of this hypothesis and potentially also improvement on germline scores for PCa aggressiveness.

A strength of the present study was the use of large, diverse datasets. Men of African ancestry are both more likely to develop metastatic PCa and more likely to have high PHS601^14,15^. Looking specifically at those with high PHS601, PRSagg/psa group based on PRSagg and PRS_PSA_ stratified both European-ancestry and African-ancestry individuals for risk of PCa metastasis (Fig 3), demonstrating further refinement of genetic risk prediction than was previously possible.

In multivariable models, we observed higher risk of aggressive PCa for men with non-European ancestry. It is well known that Black men are more likely to develop metastatic or fatal PCa. What is unclear is how much of this is attributable to inherited biological factors versus shared environmental exposures and social determinants of health that can have tremendous impact on health outcomes^29–33^. Associations found here could arise from any of these. Interestingly, our models also found an increased risk of aggressive PCa among men with East Asian ancestry, which is less common in the literature^12–14^. This association was seen in both MVP and PRACTICAL datasets, but it may only emerge when accounting for PRS_PSA_ and PRSagg and assessing for PCa aggressiveness. Asian-ancestry individuals make up a minority of the present study; further investigation in larger datasets is warranted.

Several SNPs from our PRSagg score map to genes associated with PCa risk. We found multiple SNPs (rs11665748, rs2659124, rs266863, rs1444646431) that map to the *KLK3* / *KLK2* cluster, notable because *KLK3* encodes PSA^34^. Another SNP (rs10069690) maps to telomerase reverse transcriptase (*TERT*), one of the most well-established pan-cancer susceptibility loci^35^. Other genes of interest include *IRX4* (rs386684494), a known prostate cancer risk gene regulating androgen signaling^36^, *LMTK2* (rs11763970), directly implicated in androgen receptor signaling and prostate cancer risk^37^, and *PCAT1* (rs73351629), a prostate cancer-specific oncogenic lncRNA^38^. Further exploratory biological results are described in the Supplementary Material.

Several tools are emerging for prognostication for patients on AS. These include clinical risk models^39,40^, molecular tests on biopsy specimens^41,41,42^, and digital pathology algorithms^43,44^. MRI is increasingly used to select patients for active surveillance and to monitor them during surveillance^4,45–47^. How germline features may interact with each of these is worthy of study. Given the very strong association of cribriform morphology with AS outcomes, an interesting future question will be the relationship of germline features to development of somatic cribriform phenotypes^4,28,48^. The subsets of ProtecT participants with known cribriform status and with available germline genotyping, respectively, are only partially overlapping at present. Ongoing efforts to sequence germline DNA from the full cohort will facilitate future investigation.

One limitation of this study is the use of a conservative management cohort (MVP Monitored PCa Cohort) in place of a true AS cohort. AS status was unavailable in MVP. Therefore, we approximated an AS cohort by including patients having known PCa diagnosis, first recorded treatment more than one year after diagnosis, and grade group 1 or 2 disease. Additionally, polygenic scores do not incorporate rare pathogenic variants, structural variation, epigenetic regulation, or germline–somatic interactions that can contribute to inherited architecture of PCa aggressiveness. The SNP pre-selection and LASSO approach may introduce some degree of overfitting or instability, for which .632 bootstrapping was intended to mitigate. PCa aggressiveness is largely defined by grade group at diagnosis, which is biopsy-based and subject to sampling variability in the biopsy procedure. The number of metastatic events in ProtecT was relatively small; larger cohorts are needed for further validation, given the rarity of such events in early-stage PCa.

Taken together, common germline markers contribute to a polygenic score (PRSagg) associated with PCa aggressiveness. PRSagg was independently associated with higher grade group when accounting for age, genetic ancestry, polygenic predisposition to benign elevation of PSA, and polygenic risk of any PCa. Moreover, PRSagg was associated with clinically meaningful outcomes among patients managed with active monitoring. The combination of PRSagg and PRS_PSA_ stratified men who developed PCa by their risk of developing aggressive disease. Given that PRSagg can be calculated decades before cancer develops, these results also hint at potential biological insights into the influence of the germline landscape on tumorigenesis and tumor evolution. Next steps include investigating possible biological mechanisms for germline association with somatic phenotype and evaluating clinical utility of polygenic scores for cancer aggressiveness in larger prospective datasets.

## Supporting information

Supplementary authors and affiliations

Supplementary funding and acknowledgements

## Data availability

Full summary statistics relating to the Million Veteran Program (MVP) studies are available at dbGAP accession phs001672. Summary statistics for PRACTICAL and ProtecT datasets are available upon reasonable request to.

## Acknowledgments

<u>VA Million Veteran Program Core Acknowledgements</u>

MVP Program Office

- Sumitra Muralidhar, Ph.D., Program Director US Department of Veterans Affairs, 810 Vermont Avenue NW, Washington, DC 20420
- Jennifer Moser, Ph.D., Associate Director, Scientific Programs US Department of Veterans Affairs, 810 Vermont Avenue NW, Washington, DC 20420
- Jennifer E. Deen, B.S., Associate Director, Cohort & Public Relations US Department of Veterans Affairs, 810 Vermont Avenue NW, Washington, DC 20420

MVP Steering Committee

- Co-Chair: Philip S. Tsao, Ph.D. VA Palo Alto Health Care System, 3801 Miranda Avenue, Palo Alto, CA 94304
- Co-Chair: Sumitra Muralidhar, Ph.D. US Department of Veterans Affairs, 810 Vermont Avenue NW, Washington, DC 20420
- J. Michael Gaziano, M.D., M.P.H. VA Boston Healthcare System, 150 S. Huntington Avenue, Boston, MA 02130
- Adriana Hung, M.D., M.P.H., VA Tennessee Valley Healthcare System, 1310 24th Avenue, South Nashville, TN 37212
- Dave Oslin, M.D. Philadelphia VA Medical Center, 3900 Woodland Avenue, Philadelphia, PA 19104
- Deepak Voora, M.D. Durham VA Medical Center, 508 Fulton Street, Durham, NC 27705

MVP Co-Principal Investigators

- J. Michael Gaziano, M.D., M.P.H. VA Boston Healthcare System, 150 S. Huntington Avenue, Boston, MA 02130
- Philip S. Tsao, Ph.D. VA Palo Alto Health Care System, 3801 Miranda Avenue, Palo Alto, CA 94304

MVP Core Operations

- Jessica V. Brewer, M.P.H., Director, MVP Cohort Operations VA Boston Healthcare System, 150 S. Huntington Avenue, Boston, MA 02130
- Mary T. Brophy M.D., M.P.H., Director, VA Central Biorepository VA Boston Healthcare System, 150 S. Huntington Avenue, Boston, MA 02130
- Kelly Cho, M.P.H, Ph.D., Director, MVP Phenomics VA Boston Healthcare System, 150 S. Huntington Avenue, Boston, MA 02130
- Lori Churby, B.S., Director, MVP Regulatory Affairs VA Palo Alto Health Care System, 3801 Miranda Avenue, Palo Alto, CA 94304
- Jacob T. Kean, Ph.D., Acting Director, VA Informatics and Computing Infrastructure (VINCI) VA Salt Lake City Health Care System, 500 Foothill Drive, Salt Lake City, UT 84148
- Saiju Pyarajan Ph.D., Director, Data and Computational Sciences VA Boston Healthcare System, 150 S. Huntington Avenue, Boston, MA 02130
- Robert Ringer, Pharm.D., Director, VA Albuquerque Central Biorepository New Mexico VA Health Care System, 1501 San Pedro Drive SE, Albuquerque, NM 87108
- Luis E. Selva, Ph.D., Director, MVP Biorepository Coordination VA Boston Healthcare System, 150 S. Huntington Avenue, Boston, MA 02130
- Shahpoor (Alex) Shayan, M.S., Director, MVP PRE Informatics VA Boston Healthcare System, 150 S. Huntington Avenue, Boston, MA 02130
- Brady Stephens, M.S., Principal Investigator, MVP Information Center Canandaigua VA Medical Center, 400 Fort Hill Avenue, Canandaigua, NY 14424
- Stacey B. Whitbourne, Ph.D., Director, MVP Cohort Development and Management VA Boston Healthcare System, 150 S. Huntington Avenue, Boston, MA 02130

This research is based on data from the Million Veteran Program, Office of Research and Development, Veterans Health Administration, and was supported by MVP000, MVP084 as well as award # I01 CX002635. This publication does not represent the views of the Department of Veteran Affairs or the United States Government.

This research was supported in part by the Intramural Research Program of the National Institutes of Health (NIH). The contributions of the NIH author(s) are considered Works of the United States Government. The findings and conclusions presented in this paper are those of the author(s) and do not necessarily reflect the views of the NIH or the U.S. Department of Health and Human Services.

Additional funding and acknowledgements are included in supplementary funding and acknowledgements.

