## Supplementary figures and images for "Germline polygenic score for prostate cancer aggressiveness"

### Supplementary Figure 1

#### Slide 1
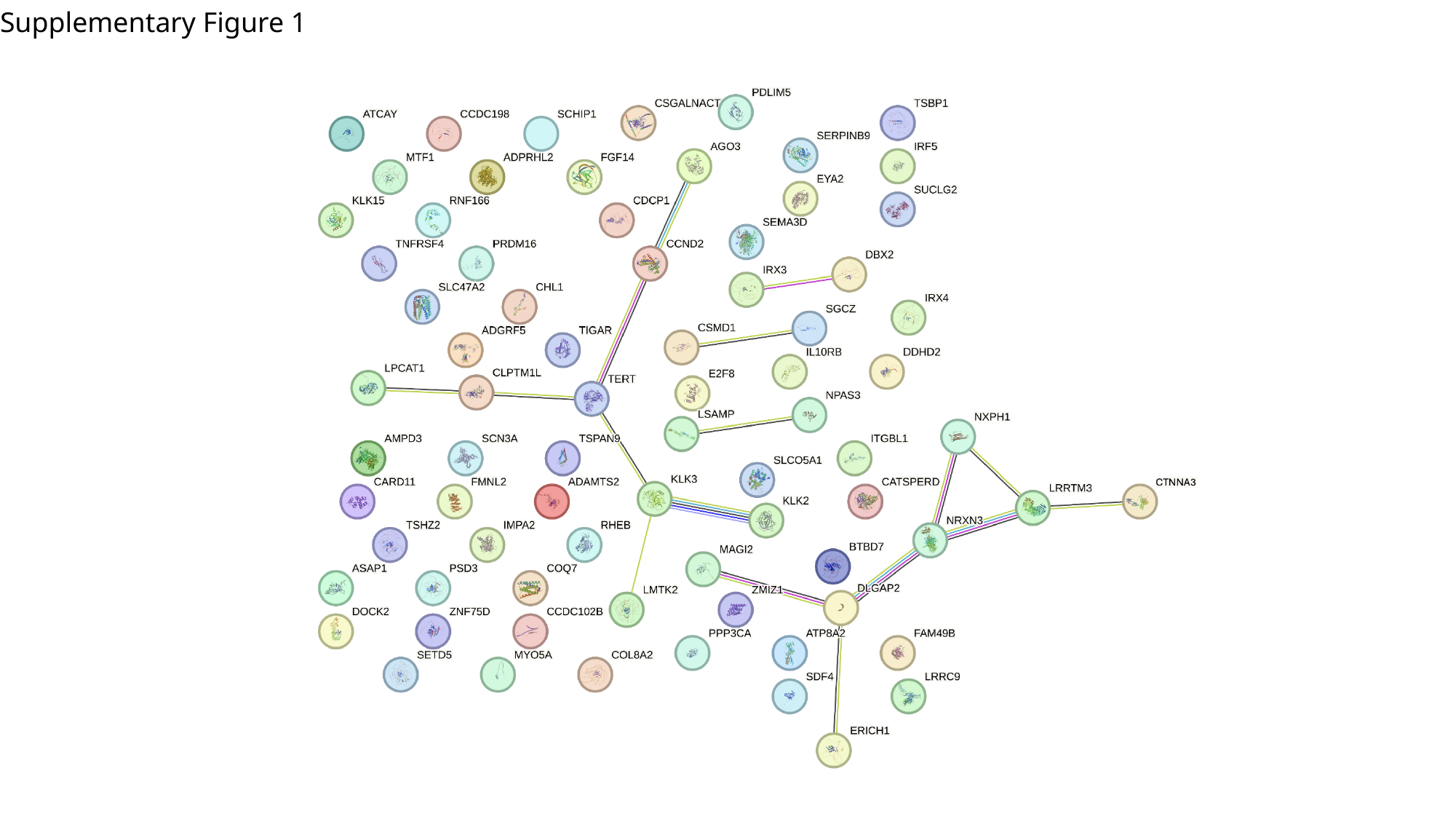

Supplementary Figure 1
