## Supplementary Material for "Germline polygenic score for prostate cancer aggressiveness"

**Proportional odds assumption testing**

To test the proportional odds assumption model, we fit a series of logistic models using different cut-points of Grade Group (Grade Group > 1, Grade Group > 2, etc.) as the outcome variable, and PRSagg as the predictor variable. The coefficient for PRSagg from these logistic models ranged from 0.40 to 0.54, whereas the coefficient from the full proportional odds model was 0.44. This suggests that the proportional odds assumption is a good approximation of the underlying data.

**STRING pathway analysis**

Using the STRING database, we conducted a pathway analysis for the set of identified genes that contain or are proximal to our PRSagg set of variants. Interactions identified by the pathway analysis from the STRING database are shown in Supplementary Table 6 and are graphically depicted in Supplementary Figure 1. This analysis identified many nodes connecting these genes, with strong connections such as *KLK2*-*KLK3* and *CLPTM1L*-*TERT* highlighting pathways related to prostate function and telomere maintenance.
