## Supplementary authors and affiliations for "Germline polygenic score for prostate cancer aggressiveness"

Artitaya Lophatananon^1^, Kenneth R Muir^1^, Teuvo LJ Tammela^2,3^, Anssi Auvinen^4^, Alison M Dunning^5^, Suzanne Chambers^6,7^, Lisa Horvath^8,9^, Wayne Tilley^10^, Gail P Risbridger^11,12^, Achala Shenali Fernando Vitharanage^13,14^, Judith A Clements^15,16,14^, Markus Aly^17,18,19^, Tobias Nordström^20,21^, Andreas Røder^22^, Stig E Bojesen^23,24^, Hein V Stroomberg^22^, Phyllis J Goodman^25^, Ian M Thompson Jr^26^, Melissa C Southey^27,28,29^, Graham G Giles^28,30,27^, Roger L Milne^28,30,27^, Niclas Håkansson^31^, Stephanie J Weinstein^32^, Janet L Stanford^33,34^, Edward Giovannucci^35^, Peter Kraft^36^, Bettina F Drake^37^, Géraldine Cancel-Tassin^38,39^, Stephen Chanock^32^, Gerald L Andriole^40^, Robert N Hoover^32^, Mitchell J Machiela^32^, Laura E Beane Freeman^32^, Michael Borre^41,42^, Dominika Wokołorczyk^43^, Jan Lubinski^43^, Thérèse Truong^44^, Hui-Yi Lin^45^, Manuel Luedeke^46^, Thomas Schnoeller^47^, Neil E Fleshner^48^, Antonio Finelli^49^, Sarah L Kerns^50^, Harry Ostrer^51^, Antonio Gómez-Caamaño^52^, Laura Fachal^53,54,55,56^, Jack A Taylor^57,58^, Jeannette T Bensen^59,60^, James L Mohler^61,60^, Eboneé Butler^59^, Trinidad Dierssen-Sotos^62,63^, Gemma Castaño-Vinyals^64,65,66,63^, Antonio Alcaraz^67^, Robert Szulkin^20,68^, Martin Eklund^20^, Meir Stampfer^69^, Sara Lindström^70^, Paula Paulo^71^, Andreia Brandão^71^, Pascal Blanchet^72^, Laurent Brureau^72^, Bernd Holleczek^73^, Ben Schöttker^74^, Sigrid Carlsson^74,75^, Chavdar Slavov^76^, Vanio Mitev^77^, Yuan Chun Ding^78^, Linda Steele^78^, Gert De Meerleer^79^, Jasmine Lim^80^, Soo-Hwang Teo^81^, Davor Lessel^82^, Tomislav Kulis^83^, Matthew Parliament^84,85^, Aswin Abraham^84,85^, Steven Joniau^86^, Manuela Gago-Dominguez^87^, Maria Elena Martinez^88^, Jennifer Cullen^89^, Maureen Sanderson^90^, Christopher A Haiman^91^, Fredrick R Schumacher^89,92^, Sara Benlloch^93,94^, Ali Amin Al Olama^93,95^, David V Conti^91^, Ying Wang^96^, Catharine ML West^97^, Eli Marie Grindedal^98^, Sue Ann Ingles^99^, Lisa F Newcomb^33,100^, Monique J Roobol^101^, Lisa Cannon-Albright^102,103^, Hardev Pandha^104^, Stephen N Thibodeau^105^, David J Hunter^106^, Elio Riboli^107^

^1^Division of Population Health, Health Services Research and Primary Care, University of Manchester, Oxford Road, Manchester, M13 9PL, UK

^2^Department of Urology, Tampere University Hospital, FI-33521 Tampere, Finland

^3^Faculty of Medicine and Health Technology, Tampere University, FI-33100 Tampere, Finland

^4^Unit of Health Sciences, Faculty of Social Sciences, Tampere University, Tampere, Finland

^5^Centre for Cancer Genetic Epidemiology, Department of Oncology, University of Cambridge, Strangeways Laboratory, Worts Causeway, Cambridge, CB1 8RN, UK

^6^Faculty of Health Sciences, Australian Catholic University

^7^Cancer Council Queensland, Fortitude Valley, QLD 4006, Australia

^8^Chris O'Brien Lifehouse (COBLH), Camperdown, Sydney, NSW 2010, Australia

^9^Garvan Institute of Medical Research, Sydney NSW 2010, Australia

^10^Dame Roma Mitchell Cancer Research Laboratories, University of Adelaide, Adelaide, South Australia, Australia

^11^Department of Anatomy and Developmental Biology, Biomedicine Discovery Institute, Monash University, Melbourne,Victoria 3800, Australia

^12^Prostate Cancer Translational Research Program, Cancer Research Division, Peter MacCallum Cancer Centre, Melbourne, VIC 3000, Australia

^13^Centre for Genomics and Personalised Health, Queensland University of Technology, Brisbane, Australia

^14^Translational Research Institute, Brisbane, Queensland 4102, Australia

^15^School of Biomedical Sciences, Faculty of Health, Queensland University of Technology, Brisbane, Queensland, Australia

^16^Centre for Genomics and Personalised Health, Queensland University of Technology, Brisbane, Queensland, Australia

^17^Department of Medical Epidemiology and Biostatistics, Karolinska Institutet, Stockholm, Sweden

^18^Department of Molecular Medicine and Surgery, Karolinska Institutet

^19^Department of Urology, Karolinska University Hospital, Solna, 171 76 Stockholm

^20^Department of Medical Epidemiology and Biostatistics, Karolinska Institutet, Stockholm, Sweden

^21^Department of Clinical Sciences at Danderyds Hospital, Karolinska Institutet, Stockholm, Sweden

^22^Copenhagen Prostate Cancer Center, Department of Urology, Rigshospitalet, Copenhagen University Hospital, DK-2730 Herlev, Copenhagen, Denmark

^23^Faculty of Health and Medical Sciences, University of Copenhagen, 2200 Copenhagen, Denmark

^24^Department of Clinical Biochemistry, Herlev and Gentofte Hospital, Copenhagen University Hospital, Herlev, 2200 Copenhagen, Denmark

^25^SWOG Statistical Center, Fred Hutchinson Cancer Research Center, Seattle, WA, USA

^26^CHRISTUS Santa Rosa Hospital – Medical Center, San Antonio, TX, USA

^27^Precision Medicine, School of Clinical Sciences at Monash Health, Monash University, Clayton, Victoria 3168, Australia

^28^Cancer Epidemiology Division, Cancer Council Victoria, 200 Victoria Parade, East Melbourne, VIC, 3002, Australia

^29^Department of Clinical Pathology, The Melbourne Medical School, The University of Melbourne, Melbourne, VIC 3010, Australia.

^30^Centre for Epidemiology and Biostatistics, Melbourne School of Population and Global Health, The University of Melbourne, Grattan Street, Parkville, VIC 3010, Australia

^31^Unit of Cardiovascular and Nutritional Epidemiology, Institute of Environmental Medicine, Karolinska Institutet, SE-171 77 Stockholm, Sweden

^32^Division of Cancer Epidemiology and Genetics, National Cancer Institute, NIH, Bethesda, Maryland, 20892, USA

^33^Division of Public Health Sciences, Fred Hutchinson Cancer Research Center, Seattle, Washington, 98109-1024, USA

^34^Department of Epidemiology, School of Public Health, University of Washington, Seattle, Washington 98195, USA

^35^Department of Epidemiology, Harvard School of Public Health, Boston, MA 02115, USA

^36^Program in Genetic Epidemiology and Statistical Genetics, Department of Epidemiology, Harvard T. H. Chan School of Public Health, Boston 02115, USA

^37^Washington University School of Medicine, 660 S. Euclid Avenue, Campus Box 8242, St. Louis, MO 63110 , USA

^38^CeRePP, Tenon Hospital, F-75020 Paris, France.

^39^Sorbonne Universite, GRC n°5 , AP-HP, Tenon Hospital, 4 rue de la Chine, F-75020 Paris, France

^40^The Washington University School of Medicine, 660 S. Euclid Avenue, Campus Box 8242, St. Louis, MO 63110 , USA

^41^Department of Urology, Aarhus University Hospital, Palle Juul-Jensen Boulevard 99, 8200 Aarhus N, Denmark

^42^Department of Clinical Medicine, Aarhus University, DK-8200 Aarhus N

^43^International Hereditary Cancer Center, Department of Genetics and Pathology, Pomeranian Medical University, 70-115 Szczecin, Poland

^44^Exposome and Heredity, CESP (UMR 1018), Faculté de Médecine, Université Paris-Saclay, Inserm, Gustave Roussy, Villejuif

^45^School of Public Health, Louisiana State University Health Sciences Center, New Orleans, LA 70112, USA

^46^genetikum, Wegenerstr. 15, D-89231 Neu-Ulm, Germany

^47^Department of Urology, University Hospital Ulm, Germany

^48^Dept. of Surgical Oncology, UHN Princess Margaret Cancer Centre, Toronto ON M5G 2M9, Canada

^49^Division of Urology, Princess Margaret Cancer Centre, Toronto ON M5G 2M9, Canada

^50^The Medical College of Wisconsin, 8701 Watertown Plank Rd., Milwaukee, WI 53226, USA

^51^Department of Pathology, Albert Einstein College of Medicine, 1300 Morris Park Avenue, Ullman 817, Bronx, NY 10461, USA

^52^Department of Radiation Oncology, Complexo Hospitalario Universitario de Santiago, SERGAS, Santiago de Compostela 15706, Spain

^53^Centre for Cancer Genetic Epidemiology, Department of Public Health and Primary Care, University of Cambridge, Strangeways Research Laboratory, Cambridge, CB2 0SR, UK

^54^Fundación Pública Galega Medicina Xenómica, Santiago De Compostela, 15706, Spain.

^55^Instituto de Investigación Sanitaria de Santiago de Compostela, Santiago De Compostela, 15706, Spain.

^56^Centro de Investigación en Red de Enfermedades Raras (CIBERER), Spain

^57^Epidemiology Branch, National Institute of Environmental Health Sciences, Research Triangle Park, NC, USA

^58^Epigenetic and Stem Cell Biology Laboratory, National Institute of Environmental Health Sciences, Research Triangle Park, NC

^59^Department of Epidemiology, University of North Carolina at Chapel Hill, Chapel Hill, NC, USA

^60^Lineberger Comprehensive Cancer Center, University of North Carolina at Chapel Hill, Chapel Hill, NC, USA

^61^Department of Urology, Roswell Park Comprehensive Cancer Center, Buffalo, NY, USA

^62^University of Cantabria-IDIVAL, Santander, Spain

^63^CIBER Epidemiología y Salud Pública (CIBERESP), 28029 Madrid, Spain

^64^ISGlobal, Barcelona, Spain

^65^IMIM (Hospital del Mar Medical Research Institute), Barcelona, Spain

^66^Universitat Pompeu Fabra (UPF), Barcelona, Spain

^67^Department and Laboratory of Urology. Hospital Clínic. Institut d’Investigacions Biomèdiques August Pi i Sunyer (IDIBAPS), Universitat de Barcelona. Spain. C/Villarroel 170; 08036 Barcelona, Spain

^68^SDS Life Science, Danderyd, Sweden

^69^Channing Division of Network Medicine, Department of Medicine, Brigham and Women's Hospital/Harvard Medical School, Boston, MA 02184, USA

^70^Department of Epidemiology, Health Sciences Building, University of Washington

^71^Cancer Genetics Group, IPO Porto Research Center (CI-IPOP) / RISE@CI-IPOP (Health Research Network), Portuguese Oncology Institute of Porto (IPO Porto) / Porto Comprehensive Cancer Center, Porto, Portugal

^72^CHU de Pointe-à-Pitre, Univ Antilles, Univ Rennes, Inserm, EHESP, Irset (Institut de recherche en santé, environnement et travail) - UMR_S 1085, Pointe-à-Pitre, France

^73^Saarland Cancer Registry, 66119 Saarbrücken, Germany

^74^Division of Clinical Epidemiology of Early Cancer Detection, German Cancer Research Center (DKFZ), D-69120, Heidelberg, Germany

^75^Department of Translational Medicine, Division of Urological Cancers, Lund University, Lund, Sweden

^76^Department of Urology and Alexandrovska University Hospital, Medical University of Sofia, 1431 Sofia, Bulgaria

^77^Molecular Medicine Center, Department of Medical Chemistry and Biochemistry, Medical University of Sofia, Sofia, 2 Zdrave Str., 1431 Sofia, Bulgaria

^78^Department of Population Sciences, Beckman Research Institute of the City of Hope, 1500 East Duarte Road, Duarte, CA 91010

^79^Ghent University, B-9000, Gent

^80^Department of Surgery, Faculty of Medicine, University of Malaya, 50603 Kuala Lumpur, Malaysia

^81^Cancer Research Malaysia (CRM), Outpatient Centre, Subang Jaya Medical Centre, Subang Jaya, Selangor, Malaysia

^82^Institute of Human Genetics, University of Regensburg, and Institute of Clinical Human Genetics, University Hospital Regensburg, Franz-Josef-Strauss-Allee 11, D-93053 Regensburg

^83^Department of Urology, University Hospital Center Zagreb, University of Zagreb School of Medicine , Zagreb, Croatia

^84^Department of Oncology, Cross Cancer Institute, University of Alberta, 11560 University Avenue, Edmonton, Alberta, Canada T6G 1Z2

^85^Division of Radiation Oncology, Cross Cancer Institute, 11560 University Avenue, Edmonton, Alberta, Canada T6G 1Z2

^86^Department of Urology, University Hospitals Leuven, Herestraat 49, Box 7003 41, BE-3000 Leuven, Belgium

^87^Health Research Institute of Santiago de Compostela Foundation (FIDIS), Cancer Genetics and Epidemiology Group, Santiago de Compostela, Spain

^88^University of California San Diego, Moores Cancer Center, Department of Family Medicine and Public Health, University of California San Diego, La Jolla, CA 92093-0012, USA

^89^Department of Population and Quantitative Health Sciences, Case Western Reserve University, Cleveland, OH 44106-7219, USA

^90^Department of Family and Community Medicine, Meharry Medical College, 1005 Dr. DB Todd Jr. Blvd., Nashville, TN 37208 USA

^91^Center for Genetic Epidemiology, Department of Preventive Medicine, Keck School of Medicine, University of Southern California/Norris Comprehensive Cancer Center, Los Angeles, CA 90015, USA

^92^Seidman Cancer Center, University Hospitals, Cleveland, OH 44106, USA.

^93^Centre for Cancer Genetic Epidemiology, Department of Public Health and Primary Care, University of Cambridge, Strangeways Research Laboratory, Cambridge CB1 8RN, UK

^94^The Institute of Cancer Research, London, SM2 5NG, UK

^95^University of Cambridge, Department of Clinical Neurosciences, Stroke Research Group, R3, Box 83, Cambridge Biomedical Campus, Cambridge CB2 0QQ, UK

^96^Department of Population Science, American Cancer Society, 250 Williams Street, Atlanta, GA 30303, USA

^97^Division of Cancer Sciences, University of Manchester, Manchester Academic Health Science Centre, Radiotherapy Related Research, The Christie Hospital NHS Foundation Trust, Manchester, M13 9PL UK

^98^Department of Medical Genetics, Oslo University Hospital, 0424 Oslo, Norway

^99^Department of Preventive Medicine, Keck School of Medicine, University of Southern California/Norris Comprehensive Cancer Center, Los Angeles, CA 90015, USA

^100^Department of Urology, University of Washington, 1959 NE Pacific Street, Box 356510, Seattle, WA 98195, USA

^101^Department of Urology, Erasmus University Medical Center, Cancer Institute, 3015 GD Rotterdam, The Netherlands

^102^Division of Epidemiology, Department of Internal Medicine, University of Utah School of Medicine, Salt Lake City, Utah 84132, USA

^103^George E. Wahlen Department of Veterans Affairs Medical Center, Salt Lake City, Utah 84148, USA

^104^The University of Surrey, Guildford, Surrey, GU2 7XH, UK

^105^Department of Laboratory Medicine and Pathology, Mayo Clinic, Rochester, MN 55905, USA

^106^Nuffield Department of Population Health, University of Oxford, United Kingdom

^107^Department of Epidemiology and Biostatistics, School of Public Health, Imperial College London, SW7 2AZ, UK
