## Supplementary funding and acknowledgements for "Germline polygenic score for prostate cancer aggressiveness"

**Supplementary Acknowledgements & Funding**

CRUK and PRACTICAL consortium

This work was supported by the Canadian Institutes of Health Research, European Commission's Seventh Framework Programme grant agreement n° 223175 (HEALTH-F2-2009-223175), Cancer Research UK Grants C5047/A7357, C1287/A10118, C1287/A16563, C5047/A3354, C5047/A10692, C16913/A6135, and The National Institute of Health (NIH) Cancer Post-Cancer GWAS initiative grant: No. 1 U19 CA 148537-01 (the GAME-ON initiative).

We would also like to thank the following for funding support: The Institute of Cancer Research and The Everyman Campaign, The Prostate Cancer Research Foundation, Prostate Research Campaign UK (now Prostate Action), The Orchid Cancer Appeal, The National Cancer Research Network UK, The National Cancer Research Institute (NCRI) UK. We are grateful for support of NIHR funding to the NIHR Biomedical Research Centre at The Institute of Cancer Research and The Royal Marsden NHS Foundation Trust.

The Prostate Cancer Program of Cancer Council Victoria also acknowledge grant support from The National Health and Medical Research Council, Australia (126402, 209057, 251533, 396414, 450104, 504700, 504702, 504715, 623204, 940394, 614296,), VicHealth, Cancer Council Victoria, The Prostate Cancer Foundation of Australia, The Whitten Foundation, PricewaterhouseCoopers, and Tattersall’s. EAO, DMK, and EMK acknowledge the Intramural Program of the National Human Genome Research Institute for their support.

Genotyping of the OncoArray was funded by the US National Institutes of Health (NIH) [U19 CA 148537 for ELucidating Loci Involved in Prostate cancer SuscEptibility (ELLIPSE) project and X01HG007492 to the Center for Inherited Disease Research (CIDR) under contract number HHSN268201200008I].

This study would not have been possible without the contributions of the following: Coordination team, bioinformatician and genotyping centers: Genotyping at CCGE, Cambridge: Caroline Baines and Don Conroy

Funding for the iCOGS infrastructure came from: the European Community's Seventh Framework Programme under grant agreement n° 223175 (HEALTH-F2-2009-223175) (COGS), Cancer Research UK (C1287/A10118, C1287/A 10710, C12292/A11174, C1281/A12014, C5047/A8384, C5047/A15007, C5047/A10692, C8197/A16565), the National Institutes of Health (CA128978) and Post-Cancer GWAS initiative (1U19 CA148537, 1U19 CA148065 and 1U19 CA148112 - the GAME-ON initiative), the Department of Defence (W81XWH-10-1-0341), the Canadian Institutes of Health Research (CIHR) for the CIHR Team in Familial Risks of Breast Cancer, Komen Foundation for the Cure, the Breast Cancer Research Foundation, and the Ovarian Cancer Research Fund.

This study would not have been possible without the contributions of the following: Per Hall (COGS); Douglas F. Easton, Paul Pharoah, Kyriaki Michailidou, Manjeet K. Bolla, Qin Wang (BCAC), Andrew Berchuck (OCAC), Rosalind A. Eeles, Douglas F. Easton, Ali Amin Al Olama, Zsofia Kote-Jarai, Sara Benlloch (PRACTICAL), Georgia Chenevix-Trench, Antonis Antoniou, Lesley McGuffog, Fergus Couch and Ken Offit (CIMBA), Joe Dennis, Alison M. Dunning, Andrew Lee, and Ed Dicks, Craig Luccarini and the staff of the Centre for Genetic Epidemiology Laboratory, Javier Benitez, Anna Gonzalez-Neira and the staff of the CNIO genotyping unit, Jacques Simard and Daniel C. Tessier, Francois Bacot, Daniel Vincent, Sylvie LaBoissière and Frederic Robidoux and the staff of the McGill University and Génome Québec Innovation Centre, Stig E. Bojesen, Sune F. Nielsen, Borge G. Nordestgaard, and the staff of the Copenhagen DNA laboratory, and Julie M. Cunningham, Sharon A. Windebank, Christopher A. Hilker, Jeffrey Meyer and the staff of Mayo Clinic Genotyping Core Facility

**Additional funding and acknowledgments from studies in PRACTICAL:**

Information of the PRACTICAL consortium can be found at <https://practical.icr.ac.uk/>

Aarhus

This study was supported by Innovation Fund Denmark, the Danish Cancer Society and The Velux Foundation (Veluxfonden).

The Danish Cancer Biobank (DCB) is acknowledged for biological material.

AHS

This work was supported by the Intramural Research Program of the NIH, National Cancer Institute, Division of Cancer Epidemiology and Genetics (Z01CP010119).

ATBC

This research was supported by the Intramural Research Program of the National Institutes of Health (NIH). The contributions of the NIH author(s) are considered Works of the United States Government. The findings and conclusions presented in this paper are those of the author(s) and do not necessarily reflect the views of the NIH or the U.S. Department of Health and Human Services.

CCI

This work was awarded by Prostate Cancer Canada and is proudly funded by the Movember Foundation - Grant # D2013-36.

The CCI group would like to thank David Murray, Razmik Mirzayans, and April Scott for their contribution to this work.

CHIPGECS

Orchid, Wuxi Second Hospital Research Funds, National Natural Science foundation of China for funding support to H Zhang (Grant No: 30671793 and 81072377), N Feng (Grant No: 81272831), SC Zhao (Grant No: 81072092 and 81328017), Liaoning Natural Science Foundation 2017, China, Item Number: 20170540536 to Zan Sun Sun and Yong-Jie Lu.

This work was conducted on behalf of the CHIPGECS Consortia. We acknowledge the contribution of doctors, nurses and postgraduate research students at the CHIPGENCS sample collecting centers.

COH

SLN is partially supported by the Morris and Horowitz Families Endowed Professorship

COSM

COSM is part of the Swedish Infrastructure for Medical Population-based Life-course and Environmental Research (SIMPLER), which receives funding from the Swedish Research Council (grants VR 2017-00644, VR 2021-00160. This work was supported by the Swedish Cancer Foundation (grant CF 20 0864).

CPCS1/CPCS2

Department of Clinical Biochemistry, Herlev and Gentofte Hospital, Copenhagen University Hospital, Herlev Ringvej 75, DK-2730 Herlev, Denmark

We thank participants and staff of the Copenhagen General Population Study for their important contributions.

CPDR

Uniformed Services University for the Health Sciences HU0001-10-2-0002 (PI: David G. McLeod, MD)

EPIC

The coordination of EPIC was financially supported by the European Commission (DG-SANCO) and the International Agency for Research on Cancer. The national cohorts (that recruited male participants) are supported by Danish Cancer Society (Denmark); German Cancer Aid, German Cancer Research Center (DKFZ), Federal Ministry of Education and Research (BMBF), Deutsche Krebshilfe, Deutsches Krebsforschungszentrum and Federal Ministry of Education and Research (Germany); the Hellenic Health Foundation (Greece); Associazione Italiana per la Ricerca sul Cancro-AIRC-Italy and National Research Council (Italy); Dutch Ministry of Public Health, Welfare and Sports (VWS), Netherlands Cancer Registry (NKR), LK Research Funds, Dutch Prevention Funds, Dutch ZON (Zorg Onderzoek Nederland), World Cancer Research Fund (WCRF), Statistics Netherlands (The Netherlands); Health Research Fund (FIS), PI13/00061 to Granada; , PI13/01162 to EPIC-Murcia), Regional Governments of Andalucía, Asturias, Basque Country, Murcia and Navarra, ISCIII RETIC (RD06/0020) (Spain); Swedish Cancer Society, Swedish Research Council and County Councils of Skåne and Västerbotten (Sweden); Cancer Research UK (14136 to EPIC-Norfolk; C8221/A29017 to EPIC-Oxford), Medical Research Council

(1000143 to EPIC-Norfolk, MR/M012190/1 to EPIC-Oxford) (United Kingdom). For information on how to submit an application for gaining access to EPIC data and/or biospecimens, please follow the instructions at http://epic.iarc.fr/access/index.php

EPICAP

The EPICAP study was supported by grants from Ligue Nationale Contre le Cancer; Institut National du Cancer (INCa); Fondation ARC; Fondation de France; Agence Nationale de sécurité sanitaire de l’alimentation, de l’environnement et du travail (ANSES); Ligue départementale du Val de Marne.

The EPICAP study group would like to thank all urologists, Antoinette Anger and Hasina Randrianasolo (study monitors), Anne-Laure Astolfi, Coline Bernard, Oriane Noyer, Marie-Hélène De Campo, Sandrine Margaroline, Louise N'Diaye, Sabine Perrier-Bonnet (Clinical Research nurses)

ESTHER

The ESTHER study was supported by a grant from the Baden Württemberg Ministry of Science, Research and Arts.

The ESTHER group would like to thank Hartwig Ziegler, Sonja Wolf, Volker Hermann, Heiko Müller, Karina Dieffenbach, Katja Butterbach for valuable contributions to the study.

FHCRC

The FHCRC studies were supported by grants R01-CA080122, R01-CA056678, R01-CA082664, and R01-CA092579, and K05-CA175147 from the US National Cancer Institute, National Institutes of Health, with additional support from the Fred Hutchinson Cancer Research Center (P30-CA015704).

We thank all the individuals who participated in these studies.

Gene-PARE

The Gene-PARE study was supported by grants 1R01CA134444 from the U.S. National Institutes of Health , PC074201 and W81XWH-15-1-0680 from the Prostate Cancer Research Program of the Department of Defense and RSGT-05-200-01-CCE from the American Cancer Society. S.L.K. is supported by 1K07CA187546 from the U.S. National Cancer Institute.

HPFS

The Health Professionals Follow-up Study was supported by grants UM1CA167552, CA133891, CA141298, and P01CA055075.

We are grateful to the participants and staff of the Physicians’ Health Study and Health Professionals Follow-Up Study for their valuable contributions, as well as the following state cancer registries for their help: AL, AZ, AR, CA, CO, CT, DE, FL, GA, ID, IL, IN, IA, KY, LA, ME, MD, MA, MI, NE, NH, NJ, NY, NC, ND, OH, OK, OR, PA, RI, SC, TN, TX, VA, WA, WY.

IMPACT

The IMPACT study was funded by The Ronald and Rita McAulay Foundation, Cancer Research UK (Grant references (C5047/A21332, C5047/A13232, C5047/A17528, and PRCPJT-Nov23/100008), the BRCA Research and Cure Alliance, Cancer Australia, AICR Netherlands A10-0227, Cancer Australia and Cancer Council Tasmania, NIHR, EU Framework 6, Cancer Councils of Victoria and South Australia, Philanthropic donation to Northshore University Health System. We acknowledge support from the National Institute for Health Research (NIHR) to the Biomedical Research Centre at The Institute of Cancer Research and Royal Marsden Foundation NHS Trust.

We acknowledge the IMPACT study steering committee, collaborating centres and participants.

IPO-Porto

The IPO-Porto study was funded by Fundação para a Ciência e a Tecnologia (FCT; UIDP/0076/2020, CEECINST/00091/2018, and 2021.03835.CEECIND) and by IPO-Porto Research Center (CI-IPOP-24-2015).

We would like to express our gratitude to all patients and families who have participated in this study.

KARUPROSTATE

The Karuprostate study was supported by the French National Health Directorate, the Association pour la Recherche sur le Cancer, la Ligue Nationale contre le Cancer, the French Agency for Environmental and Occupational Health Safety (ANSES) and by the Association pour la Recherche sur les Tumeurs de la Prostate. We would like to thank Séverine Ferdinand for valuable contributions to the study.

KULEUVEN

F.C. and S.J. are holders of grants from FWO Vlaanderen (G.0684.12N and G.0830.13N), the Belgian federal government (National Cancer Plan KPC_29_023), and a Concerted Research Action of the KU Leuven (GOA/15/017). TVDB is holder of a doctoral fellowship of the FWO.

Malaysia

The study was funded by the University Malaya High Impact Research Grant (HIR/MOHE/MED/35 to A.R).

We thank all associates in the Urology Unit, University of Malaya, Cancer Research Malaysia (CRM) and the Malaysian Men's Health Initiative (MMHI).

MCC-Spain

The study was partially funded by the "Accion Transversal del Cancer", approved on the Spanish Ministry Council on the 11th October 2007, by the Instituto de Salud Carlos III-FEDER (PI08/1770, PI09/00773-Cantabria, PI11/01889-FEDER, PI12/00265, PI12/01270, PI12/00715, PI15/00069), by the Fundación Marqués de Valdecilla (API 10/09), by the Spanish Association Against Cancer (AECC) Scientific Foundation and by the Catalan Government DURSI grant 2009SGR1489. Samples: Biological samples were stored at the Parc de Salut MAR Biobank (MARBiobanc; Barcelona) which is supported by Instituto de Salud Carlos III FEDER (RD09/0076/00036). Also sample collection was supported by the Xarxa de Bancs de Tumors de Catalunya sponsored by Pla Director d'Oncologia de Catalunya (XBTC). ISGlobal acknowledges support from the Spanish Ministry of Science and Innovation through the “Centro de Excelencia Severo Ochoa 2019-2023” Program (CEX2018-000806-S), and support from the Generalitat de Catalunya through the CERCA Program.

We acknowledge the contribution from Esther Gracia-Lavedan in preparing the data. We thank all the subjects who participated in the study and all MCC-Spain collaborators.

MCCS

Melbourne Collaborative Cohort Study (MCCS) cohort recruitment was funded by VicHealth and Cancer Council Victoria. The MCCS was further augmented by Australian National Health and Medical Research Council grants 209057, 396414 and 1074383 and by infrastructure provided by Cancer Council Victoria. Cases and their vital status were ascertained through the Victorian Cancer Registry.

The Prostate Cancer Program of Cancer Council Victoria also acknowledge grant support from The National Health

and Medical Research Council, Australia (126402, 209057, 251533, 396414, 450104, 504700, 504702, 504715, 623204, 940394, 614296), VicHealth, Cancer Council Victoria, The Prostate Cancer Foundation of Australia, The Whitten Foundation, PricewaterhouseCoopers, and Tattersall’s.

MOFFITT

The Moffitt group was supported by the US National Cancer Institute (R01CA128813, PI: J.Y. Park).

NMHS

Funding for the Nashville Men's Health Study (NMHS) was provided by the National Institutes of Health Grant numbers: RO1CA121060

PCaP

The North Carolina - Louisiana Prostate Cancer Project (PCaP) and the Health Care Access and Prostate Cancer Treatment in North Carolina (HCaP-NC) study are carried out as collaborative studies supported by the Department of Defense contract DAMD 17-03-2-0052 and the American Cancer Society award RSGT-08-008-01-CPHPS, respectively.

The authors thank the staff, advisory committees and research subjects participating in the PCaP study for their important contributions.

We would like to acknowledge the UNC BioSpecimen Facility and the LSUHSC Pathology Lab for our DNA extractions, blood processing, storage and sample disbursement (https://genome.unc.edu/bsp). For studies using PCaP tissue please include: We would like to acknowledge the RPCI Department of Urology Tissue Microarray and Immunoanalysis Core for our tissue processing, storage and sample disbursement. For studies using HCaP-NC follow-up data please use: The Health Care Access and Prostate Cancer Treatment in North Carolina (HCaP-NC) study is carried out as a collaborative study supported by the American Cancer Society award RSGT-08-008-01-CPHPS. The authors thank the staff, advisory committees and research subjects participating in the HCaP-NC study for their important contributions.

PCMUS

The PCMUS study was supported by the Bulgarian National Science Fund, Ministry of Education and Science (contract DOO-119/2009; DUNK01/2-2009; DFNI-B01/28/2012) with additional support from the Science Fund of Medical University - Sofia (contract 51/2009; 8I/2009; 28/2010; ).

PHS

The Physicians’ Health Study was supported by grants CA34944, CA40360, CA097193, HL26490 and HL34595.

We are grateful to the participants and staff of the Physicians’ Health Study and Health Professionals Follow-Up Study for their valuable contributions, as well as the following state cancer registries for their help: AL, AZ, AR, CA, CO, CT, DE, FL, GA, ID, IL, IN, IA, KY, LA, ME, MD, MA, MI, NE, NH, NJ, NY, NC, ND, OH, OK, OR, PA, RI, SC, TN, TX, VA, WA, WY.

PLCO

This PLCO study was supported by the Intramural Research Program of the Division of Cancer Epidemiology and Genetics, National Cancer Institute, NIH

The authors thank Drs. Christine Berg and Philip Prorok, Division of Cancer Prevention at the National Cancer Institute, the screening center investigators and staff of the PLCO Cancer Screening Trial for their contributions to the PLCO Cancer Screening Trial. We thank Mr. Thomas Riley, Mr. Craig Williams, Mr. Matthew Moore, and Ms. Shannon Merkle at Information Management Services, Inc., for their management of the data and Ms. Barbara O’Brien and staff at Westat, Inc. for their contributions to the PLCO Cancer Screening Trial. We also thank the PLCO study participants for their contributions to making this study possible.

PRAGGA

PRAGGA was supported by Programa Grupos Emergentes, Cancer Genetics Unit, CHUVI Vigo Hospital, Instituto de Salud Carlos III, Spain.

PRAGGA wishes to thank Victor Muñoz Garzón, Manuel Enguix Castelo, Sara Miranda Ponte, Carmen M Redondo, Manuel Calaza, Francisco Gude Sampedro, Joaquín González-Carreró and the staff of the Department of Pathology and Biobank of University Hospital Complex of Vigo, Instituto de Investigación Sanitaria Galicia Sur (IISGS), SERGAS, Vigo, Spain; Máximo Fraga, José Antúnez and the Biobank of University Hospital Complex of Santiago, Santiago de Compostela, Spain; and Maria Torres, Angel Carracedo and the Galician Foundation of Genomic Medicine.

PROCAP

PROCAP was supported by the Swedish Cancer Foundation (08-708, 09-0677).

We thank and acknowledge all of the participants in the PROCAP study. We thank Carin Cavalli-Björkman and Ami Rönnberg Karlsson for their dedicated work in the collection of data. Michael Broms is acknowledged for his skilful work with the databases. KI Biobank is acknowledged for handling the samples and for DNA extraction. We acknowledge The NPCR steering group: Pär Stattin (chair), Anders Widmark, Stefan Karlsson, Magnus Törnblom, Jan Adolfsson, Anna Bill-Axelson, Ove Andrén, David Robinson, Bill Pettersson, Jonas Hugosson, Jan-Erik Damber, Ola Bratt, Göran Ahlgren, Lars Egevad, and Roy Ehrnström.

PROFILE

We would like to acknowledge the support of the Ronald and Rita McAulay Foundation, Cancer Research UK, Prostate Cancer UK and Movember. We also acknowledge support from the National Institute for Health Research (NIHR) to the Biomedical Research Centre at The Institute of Cancer Research and Royal Marsden Foundation NHS Trust

We acknowledge the Profile study steering committee and participants.

PROGReSS

This research was partially supported by funding from the Spanish Instituto de Salud Carlos III (ISCIII), an initiative of the Spanish Ministry of Economy and Innovation, co-financed by the European Regional Development Fund (ERDF/FEDER) (PI25/00744, PI22/00589, PI19/01424, PI16/00046, PI13/02030, PI10/00164, INT24/00023, INT20/00071, INT17/00133, INT16/00154, INT15/00070, and DTS24/00083). Additional support was provided by the Xunta de Galicia through the “Programa de Consolidación y Estructuración de Unidades de Investigación Competitivas GAIN” (GPC-IN607B2025/09); the Centro de Investigación Biomédica en Red de Enfermedades Raras (CIBERER; ACCI 2016: ER17P1AC7112/2018); the Mutua Madrileña Foundation (FMMA; MMA2018-1); and the Spanish Association Against Cancer (AECC; PRYES211091VEGA), through grants awarded to A. Vega. We would like to thank the patients for their contribution to the study

ProMPT/ProtecT

CRUK, NIHR, MRC, Cambridge Biomedical Research Centre

ProtecT would like to acknowledge the support of The University of Cambridge, Cancer Research UK. Cancer Research UK grants [C8197/A10123] and [C8197/A10865] supported the genotyping team. We would also like to acknowledge the support of the National Institute for Health Research which funds the Cambridge Bio-medical Research Centre, Cambridge, UK. We would also like to acknowledge the support of the National Cancer Research Prostate Cancer: Mechanisms of Progression and Treatment (PROMPT) collaborative (grant code G0500966/75466) which has funded tissue and urine collections in Cambridge. We are grateful to staff at the Welcome Trust Clinical Research Facility, Addenbrooke’s Clinical Research Centre, Cambridge, UK for their help in conducting the ProtecT study. We also acknowledge the support of the NIHR Cambridge Biomedical Research Centre, the NIHR HTA (ProtecT grant) and the NCRI / MRC (ProMPT grant) for help with the bio-repository. The UK Department of Health funded the ProtecT study through the NIHR Health Technology Assessment Programme (projects 96/20/06, 96/20/99). The ProtecT trial and its linked ProMPT and CAP (The Cluster Randomized Trial of PSA Testing for Prostate Cancer) studies are supported by Department of Health, England; Cancer Research UK grant number C522/A8649, C11043/A4286, C18281/A8145, C18281/A11326, and C18281/A15064, Medical Research Council of England grant number G0500966, ID 75466 and The NCRI, UK. The epidemiological data for ProtecT were generated though funding from the Southwest National Health Service Research and Development. DNA extraction in ProtecT was supported by USA Dept of Defense award W81XWH-04-1-0280, Yorkshire Cancer Research and Cancer Research UK. The authors would like to acknowledge the contribution of all members of the ProtecT study research group.

The views and opinions expressed therein are those of the authors and do not necessarily reflect those of the Department of Health of England. The bio-repository from ProtecT is supported by the NCRI (ProMPT) Prostate Cancer Collaborative and the Cambridge BMRC grant from NIHR.

We acknowledge support from the National Cancer Research Institute (National Institute of Health Research (NIHR) Collaborative Study: “Prostate Cancer: Mechanisms of Progression and Treatment (PROMPT)” (grant G0500966/75466).

We thank the National Institute for Health Research, Hutchison Whampoa Limited, the Human Research Tissue Bank (Addenbrooke’s Hospital), and Cancer Research UK. The authors would like to thank those men with prostate cancer and the subjects who have donated their time and their samples to the Cambridge Biorepository, which were used in this research. We also would like to acknowledge to support of the research staff in S4 who so carefully curated the samples and the follow-up data (Jo Burge, Marie Corcoran, Anne George, and Sara Stearn).

Department of Health and Social Care disclaimer: The views expressed are those of the author(s) and not necessarily those of the NHS, the NIHR or the Department of Health and Social Care.

The CAP trial was funded by grants C11043/A4286, C18281/A8145, C18281/A11326, C18281/A15064; and C18281/A24432 from Cancer Research UK. The UK Department of Health, National Institute of Health Research provided partial funding.

PROtEuS

PROtEuS was supported financially through grants from the Canadian Cancer Society [13149, 19500, 19864, 19865] and the Cancer Research Society, in partnership with the Ministère de l'enseignement supérieur, de la recherche, de la science et de la technologie du Québec, the Fonds de la recherche du Québec - Santé, the Ministère du Développement Économique, de l’Innovation et de l’Exportation du Québec and the Canada Research Chairs Program.

PROtEuS would like to thank its collaborators and research personnel, and the urologists involved in subjects recruitment. We also wish to acknowledge the special contribution made by Ann Hsing and Anand Chokkalingam to the conception of the genetic component of PROtEuS.

QLD

The QLD research is supported by The National Health and Medical Research Council (NHMRC) Australia Project Grants [390130, 1009458] and NHMRC Career Development Fellowship, Cancer Australia PdCCRS and Cancer Council Queensland funding to J Batra

The QLD team would like to acknowledge and sincerely thank the urologists, pathologists, data managers and patient participants who have generously and altruistically supported the QLD cohort.

SABOR

The SABOR research is supported by NIH/NCI Early Detection Research Network, grant U01 CA0866402-18. Also supported by the Cancer Center Support Grant to the Mays Cancer Center from the National Cancer Institute (US) P30 CA054174

SCCS

SCCS is funded by NIH grant R01 CA092447, and SCCS sample preparation was conducted at the Epidemiology Biospecimen Core Lab that is supported in part by the Vanderbilt-Ingram Cancer Center (P30 CA68485). Data on SCCS cancer cases used in this publication were provided by the Alabama Statewide Cancer Registry; Kentucky Cancer Registry, Lexington, KY; Tennessee Department of Health, Office of Cancer Surveillance; Florida Cancer Data System; North Carolina Central Cancer Registry, North Carolina Division of Public Health; Georgia Comprehensive Cancer Registry; Louisiana Tumor Registry; Mississippi Cancer Registry; South Carolina Central Cancer Registry; Virginia Department of Health, Virginia Cancer Registry; Arkansas Department of Health, Cancer Registry, 4815 W. Markham, Little Rock, AR 72205. The Arkansas Central Cancer Registry is fully funded by a grant from National Program of Cancer Registries, Centers for Disease Control and Prevention (CDC). Data on SCCS cancer cases from Mississippi were collected by the Mississippi Cancer Registry which participates in the National Program of Cancer Registries (NPCR) of the Centers for Disease Control and Prevention (CDC). The contents of this publication are solely the responsibility of the authors and do not necessarily represent the official views of the CDC or the Mississippi Cancer Registry.

SCPCS

SCPCS is funded by CDC grant S1135-19/19, and SCPCS sample preparation was conducted at the Epidemiology Biospecimen Core Lab that is supported in part by the Vanderbilt-Ingram Cancer Center (P30 CA68485).

SEARCH

SEARCH is funded by a programme grant from Cancer Research UK [C490/A10124] and supported by the UK National Institute for Health Research Biomedical Research Centre at the University of Cambridge. The University of Cambridge has received salary support in respect of PP from the NHS in the East of England through the Clinical Academic Reserve.

SFPCS

SFPCS was funded by California Cancer Research Fund grant 99-00527V-10182

SNP_Prostate_Ghent

The study was supported by the National Cancer Plan, financed by the Federal Office of Health and Social Affairs, Belgium.

SPAG

Manchester Cancer Research Centre and MRC – Medical Research Council for PhD studentship work, MUG - Male Uprising in Guernsey, Hope for Guernsey and Wessex Medical Research for research support

STHM2

STHM2 was supported by grants from The Strategic Research Programme on Cancer (StratCan), Karolinska Institutet; the Linné Centre for Breast and Prostate Cancer (CRISP, number 70867901), Karolinska Institutet; The Swedish Research Council (number K2010-70X-20430-04-3) and The Swedish Cancer Society (numbers 11-0287 and 11-0624); Stiftelsen Johanna Hagstrand och Sigfrid Linnérs minne; Swedish Council for Working Life and Social Research (FAS), number 2012-0073

The authors acknowledge the Karolinska University Laboratory, Aleris Medilab, Unilabs and the Regional Prostate Cancer Registry for performing analyses and help to retrieve data. Carin Cavalli–Björkman and Britt-Marie Hune for their enthusiastic work as research nurses. Astrid Björklund for skilful data management. We wish to thank the BBMRI.se biobank facility at Karolinska Institutet for biobank services.

SWOG-PCPT/SWOG-SELECT

PCPT is funded by Public Health Service grants U10CA37429 and 5UM1CA182883 from the National Cancer Institute.

The authors thank the site investigators and staff and, most importantly, the participants from PCPT who donated their time to this trial.

TAMPERE

The Tampere (Finland) study was supported by the Academy of Finland (251074), The Finnish Cancer Organisations, Sigrid Juselius Foundation, and the Competitive Research Funding of the Tampere University Hospital (X51003). The PSA screening samples were collected by the Finnish part of ERSPC (European Study of Screening for Prostate Cancer).

TAMPERE would like to thank Riina Liikanen, Liisa Maeaettaenen and Kirsi Talala for their work on samples and databases.

Toronto

Prostate Cancer Canada Movember Discovery Grant (D2013-17) to RJH; Canadian Cancer Society Research Institute Career Development Award in Cancer Prevention (2013-702108) to RJH

UKGPCS

UKGPCS would also like to thank the following for funding support: The Institute of Cancer Research and The Everyman Campaign, The Prostate Cancer Research Foundation, Prostate Research Campaign UK (now Prostate Cancer UK), The Orchid Cancer Appeal, The National Cancer Research Network UK, The National Cancer Research Institute (NCRI) UK. We are grateful for support of NIHR funding to the NIHR Biomedical Research Centre at The Institute of Cancer Research and The Royal Marsden NHS Foundation Trust. UKGPCS should also like to acknowledge the NCRN nurses, data managers and Consultants for their work in the UKGPCS.

UKGPCS would like to thank all urologists and other persons involved in the planning, coordination, and data collection of the study. KM and AL were in part supported from the NIHR Manchester Biomedical Research Centre

ULM

The Ulm group received funds from the German Cancer Aid (Deutsche Krebshilfe).

WUGS

WUGS would like to thank the following for funding support: The Anthony DeNovi Fund, the Donald C. McGraw Foundation, and the St. Louis Men’s Group Against Cancer.
